# Health equity impacts of climate change in the UK: a rapid systematic review and interactive evidence gap map

**DOI:** 10.1101/2025.05.14.25327602

**Authors:** Matthew L Bosworth, Zalaya Simmons, Rhiannon Cordiner, Tabitha Kavoi, Beti Evans, Tara Quinn, Barbara Rubiell, Amy Jackson, Ahimza Nagasivam, Nicola Pearce-Smith, Lea Berrang-Ford, Daphne Duval

## Abstract

**Objective:** To assess and summarise the evidence on the health equity impacts of climate change in the UK.

**Methods:** We conducted a rapid systematic review (search: February 2024) of primary studies reporting on the health impacts of climate change exposure pathways (hazards; health risks; solutions and responses) for populations at-risk of experiencing health inequalities, identified using the Core20PLUS framework (people experiencing the greatest deprivation; people from protected characteristics groups related to ethnicity, gender reassignment, sexual orientation, and religion or belief; people from inclusion health groups).

**Results:** Of the 20,014 records identified, 24 were included. Of these, 20 reported on people experiencing the greatest deprivation, with some evidence that increase in ambient temperature, extreme cold, and extreme heat had similar impacts on mortality in the most and least deprived areas. No conclusions could be drawn for this population group for other outcomes or exposures due to the limited number of studies identified and their methodological quality.

In people experiencing homelessness, 2 studies reported that increase in ambient temperature was associated with adverse health impacts but the disproportionate impact on this population group was not assessed.

No conclusions could be drawn for ethnic minority groups or people with drug and/or alcohol dependence due to the limited evidence identified. No studies were identified for the remaining population groups.

**Conclusions:** We identified important evidence gaps and methodological limitations of the existing evidence on the health equity impacts of climate change the UK that should be used to inform future research in this area.

## 1. Introduction

Climate change is a major challenge for public health worldwide. While the impacts of climate change on health are expected to be most substantial in low- and middle-income countries (1), there are also likely to be impacts in high-income countries. There is growing evidence for adverse health impacts in the UK due to multiple, interacting factors such as increasing ambient temperatures and temperature extremes, increased frequency and intensity of flooding, and increased potential for the spread of vector-borne disease (2). Impacts on health are wide-ranging and can be both direct (for example, increased risk of mortality associated with extreme heat) and indirect (such as negative mental health effects associated with displacement due to flooding). Climate change also has implications for health care delivery, impacting staff, infrastructure, supply chains, and patient mobility (3).

A combination of environmental, socioeconomic, and biological factors influence susceptibility to the health impacts of climate change by affecting the degree of exposure and sensitivity to climate change exposure pathways, and the ability to adapt to a changing climate (4). For example, lower socioeconomic status may increase exposure to climate change hazards (such as increases in temperature and flooding) or climate change related health risks (such as changes to air quality and changes in vector borne diseases) due to factors such as inadequate housing and higher occupational exposure, in turn increasing the risk of adverse health outcomes (5). In addition, certain groups may be physiologically more sensitive to the health impacts of climate change exposure pathways. For example, children are more susceptible to adverse health impacts of heatwaves (6, 7) and older adults face increased risks due to age-related declines in thermoregulation, mobility, and pre-existing chronic conditions, such as cardiovascular and respiratory diseases, which can be exacerbated by extreme heat (8, 9). Additionally, pregnancy is associated with an increased risk of heat-related complications, including pre-term birth and low birth weight, as well as heightened vulnerability to air pollution-related adverse pregnancy outcomes (10, 11). While a range of health co-benefits have been identified for climate change adaptation and mitigation measures (12), it is important to consider the potential impacts of these interventions on groups who experience inequalities to ensure that these measures do not further exacerbate these inequalities (13).

The Core20PLUS framework was developed by NHS England (14) and has been adopted by the UK Health Security Agency (UKHSA) as part of the Health Equity for Health Security Strategy 2023 to 2026 (15). In this framework, Core20 refers to the 20% most deprived of the national population as identified by the Index of Multiple Deprivation (IMD), and PLUS refers to other groups that are known to be at risk of experiencing health inequalities (such as people with protected characteristics and population groups that are socially excluded, known as inclusion health groups). As climate change will influence many of the wider determinants of health that often impact these population groups unequally, it is important to identify those populations most at risk from the health impacts of climate change to mitigate implications for health inequity.

We previously conducted a rapid mapping review on the health equity impacts of climate change in the UK. This identified, categorised, and systematically assessed the methodological quality of the included studies and mapped them accordingly into an interactive evidence gap map (16). We focused on the Core20PLUS population groups for which limited or no review-level evidence was identified in an initial scoping exercise: people experiencing the greatest deprivation, inclusion health groups, and people with protected characteristics relating to ethnicity, religion or belief, sexual orientation, and gender reassignment. The corresponding report, which did not include a narrative synthesis of the evidence identified, was published on the UK government website (17). The aim of this paper is to provide a summary of the main findings of the mapping review and to conduct and report on the narrative synthesis of the evidence identified in order to inform future research in this area.

## 2. Methods

A rapid systematic review was conducted following streamlined systematic methodologies to accelerate the review process (18). The methods for the rapid mapping review have been described in detail previously (17). The review protocol was published on the Open Science Framework before starting the review (19). Modifications to the protocol are available in Supplementary Material 1.

2.1. Eligibility criteria

The inclusion and exclusion criteria for this review are available in Supplementary Material 2. We included primary studies reporting on the health impacts of climate change in the UK for the following Core20PLUS population groups:

- people experiencing the greatest deprivation (defined in the Core20PLUS framework as the most deprived 20% of the population as identified by the IMD, although for this review income and other deprivation measures were considered for inclusion)
- people from protected characteristics groups related to ethnicity (referred to as ‘ethnic minority groups’ in this review), gender reassignment, sexual orientation, and religion or belief
- people from inclusion health groups, including: people experiencing homelessness; people with drug and/or alcohol dependence; people in contact with the criminal justice system; vulnerable migrants; Gypsy, Roma and Traveller communities; sex workers, victims of modern slavery; and other groups with experience of social exclusion

Studies conducted in settings related to these population groups (such as prisons, asylum seeker accommodation settings, rehabilitation centres or temporary housing accommodation) were also considered for inclusion. Studies conducted in the general population that reported on one of the populations of interest in subgroup analysis were included.

The remaining Core20PLUS groups (people from protected characteristics groups related to age, sex, disability, pregnancy and maternity, and people with pre-existing health conditions, including mental health conditions) were out of scope for this review as our initial scoping exercise had identified existing review-level evidence for these groups, as described in our original report (17).

The eligible climate change exposure pathways were:

- climate change related hazards (increase in ambient temperature, extreme heat, extreme cold, heavy rainfall and flooding, drought, and other extreme weather events)
- climate change related health risks (changes to vector ecology, changes to food supply and safety, changes to water supply and safety, changes to air quality, and environmental degradation)
- climate change related solutions (mitigation interventions and policy, adaptation interventions and policy, community resilience, and disaster risk reduction)

Studies reporting on climate change related health risks and solutions were only included if findings were explicitly linked to climate change. This more selective eligibility criterion was used because not all changes to air quality, for example, can be directly attributed to climate change. We did not apply this criterion for studies on climate change related hazards, such as extreme heat and flooding, because these exposure pathways can be more directly attributed to climate change.

In terms of outcomes, studies were considered for inclusion if they reported on observed or projected health effects associated with climate change exposure pathways. This included: mortality (all causes and cause-specific), morbidity (such as respiratory and cardiovascular diseases and mental health conditions), maternal and child outcomes, healthcare usage (such as Accident and Emergency (A&E) visits or 999 calls), projected health measures (such as health impact scores or life-years gained), and proximal determinants of health (such as exposure to air pollution, exposure to infectious diseases and vectors, and access to healthcare).

Primary studies (observational studies, case series and case reports, ecological studies, mixed-methods studies, qualitative studies, and modelling studies) that were published as peer-reviewed journal articles, pre-prints or grey literature between 1 January 2010 and 19 February 2024 were considered for inclusion. Articles not available in English were excluded.

### 2.2. Search strategy

Database searches (Ovid Medline, Ovid Embase, Web of Science (Science Citation Index and Social Sciences Citation Index), the Finding Accessible Inequalities Research (FAIR) database and the King’s Fund Library) were conducted on 18 July 2023 and updated on 19 February 2024. We also conducted an extensive grey literature search of 11 resources (see full list in Supplementary Material 3) on 22 August 2023.

The search strategies were drafted by an information scientist and peer-reviewed by a second information scientist (see Supplementary Material 4 for the search strategies). A validated UK geographic search filter was used for Ovid Medline and Embase to limit the evidence retrieved to UK settings (20, 21). As there is no validated UK geographic search filter available for Web of Science, we used a UK search strategy that had been adapted from the Ovid Medline filter by an information scientist.

Citation analysis (forwards and backwards analysis using citationchaser (22) and co-citation analysis using Web of Science) was conducted on 17 October 2023 using the papers included from the initial database search results (conducted on 18 July 2023) as seed papers.

We also consulted topic experts and checked reference lists of relevant systematic reviews for additional papers.

### 2.3. Screening

Results from the initial database searches were deduplicated using Deduklick (23) and imported into EPPI-Reviewer web version (24) for screening. Title and abstract screening of the original database search was completed in triplicate by 3 reviewers for 10% of the studies, with disagreements resolved by discussion among the review team. The remaining 90% was screened by one reviewer. The same approach was used for screening of the database search update, except that 2 reviewers (rather than 3) screened 10% of records in duplicate and Rayyan was used for screening (25).

Title and abstract screening of the results from the grey literature searches was completed by one reviewer using Rayyan (25) and Microsoft Excel, depending on the export format available. One reviewer completed title and abstract screening of the results of the citation analysis using EPPI-Reviewer web version (24). Checking the reference lists of relevant systematic reviews was also done by one reviewer.

Full text screening of records from all sources was done by one reviewer and checked by a second using EPPI-Reviewer web version (24), with disagreements resolved by discussion.

### 2.4. Data extraction

Data for each study were extracted into a pre-designed Microsoft Word template, which was first piloted by 2 reviewers involved in the data extraction process. Data were extracted on: study design, objective, population group, setting, study period, climate change exposure pathway, health outcomes, and relevant results.

Data extraction was conducted by one reviewer and checked by a second, with disagreements resolved by discussion. The data extraction tables are available in the supplementary materials of the mapping review (17).

### 2.5. Critical appraisal

Critical appraisal of observational studies was done in duplicate by 2 reviewers using the quality criteria checklist (QCC) for primary research (26, 27). Studies were given a quality rating of high, medium, or low, which reflects the methodological quality of a study (how well a study was conducted to minimise potential risk of bias), but does not take into account the risk of bias inherent to different study designs. Therefore, each study was classified into one of 4 study classes based on the hierarchy of evidence (26): class A (randomised controlled trials – not included in this review), class B (cohort studies), class C (for example, case-crossover studies, time series) and class D (for example, cross-sectional and before-after studies). Modelling studies were not critically appraised using a validated tool. Instead, the main methodological limitations were identified by the reviewers and summarised in the data extraction tables. See the original report for further details (17).

### 2.6. Synthesis

Visual synthesis was performed by generating an interactive evidence gap map in EPPI-Mapper (28). The framework for the evidence gap map was designed before the review started (19) and is described in detail in the original mapping review (17).

As a narrative synthesis was not provided in the original mapping review, for this paper we conducted a narrative synthesis of the findings of the included studies to summarise the available evidence by climate change exposure pathways, population groups, and health outcomes.

## 3. Results

### 3.1. Study selection

The initial database search (conducted on 18 July 2023) returned 21,559 records. After removal of duplicates, 15,325 records were screened on title and abstract. Of these, 244 full-text articles were assessed for eligibility and 19 were included in this review.

Citation searching identified a further 862 unique records (after deduplication), of which 56 full text articles were assessed for eligibility and 3 were included in this review.

The grey literature searches returned 1,555 records. After deduplication, 1,512 records were screened on title and abstract, of which 54 full text articles were sought for retrieval and 52 assessed for eligibility (2 articles could not be retrieved). No additional unique studies meeting the inclusion criteria were identified.

The database search update (conducted on 19 February 2024) returned 2,910 records. After removal of duplicates, 2,315 records were screened on title and abstract. Of these, 76 full-text articles were assessed for eligibility, and one was included in the review. Since no articles were identified by the original grey literature search, we agreed to not update these searches.

No additional unique studies were identified by searching reference lists of relevant reviews or by expert recommendation.

In total, 20,014 records were screened on title and abstract, of which 428 articles were screened on full text, resulting in 23 articles reporting on 24 studies being included in this review (one article reported on 2 studies). **Figure 1** shows the combined PRISMA flowchart; separate flowcharts for the original search and the search update can be found in the original report (17). The list of articles excluded on full text (with exclusion reasons) can be found in Supplementary Material 5.

**Figure 1.**
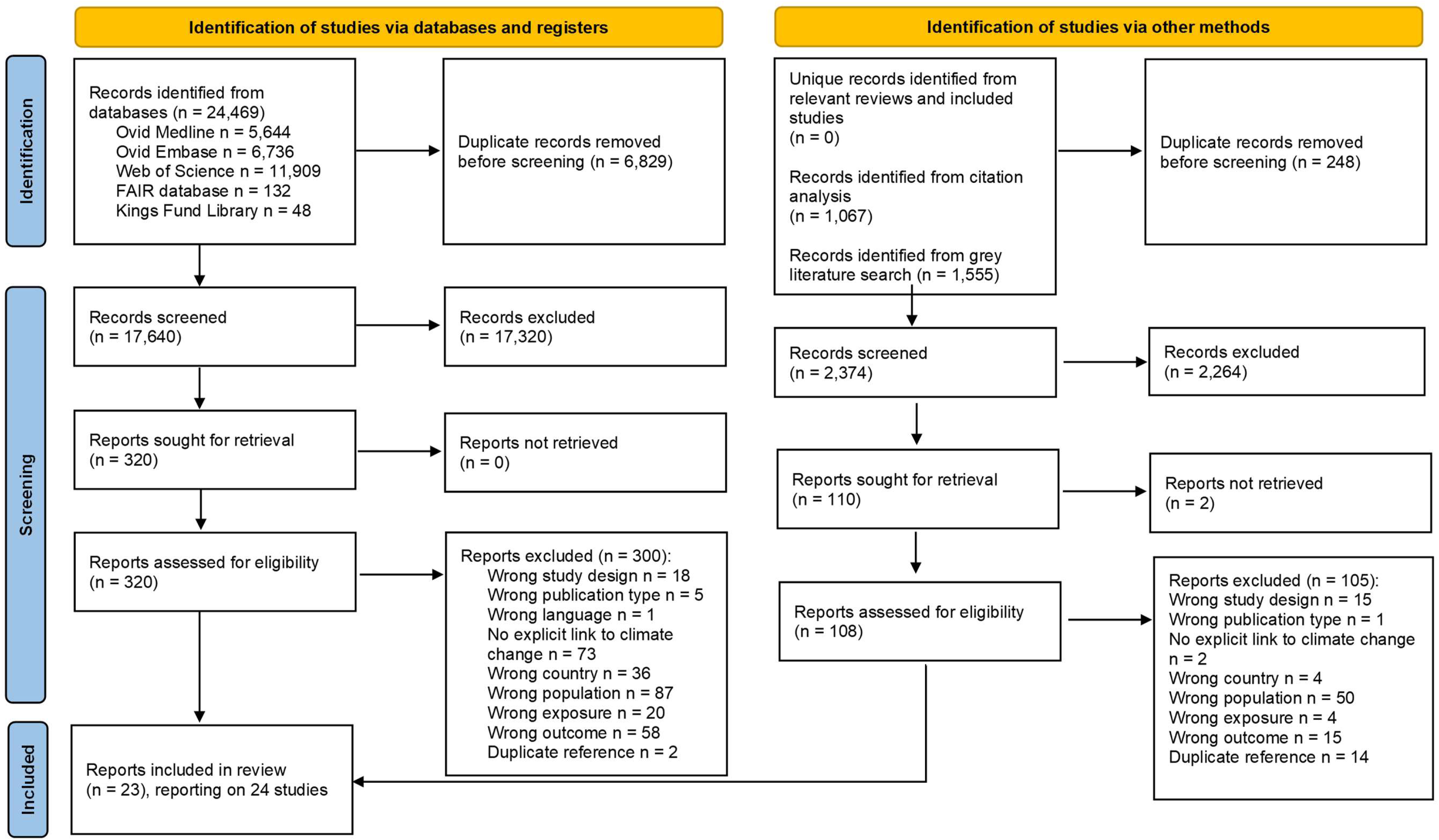
**PRISMA diagram**

### 3.2. Study characteristics

The 24 included studies were conducted across the UK: 9 studies in England (29–37), 8 studies in individual cities or regions (38–44), 4 studies in England and Wales (45–48), 2 studies in Great Britain (England, Wales and Scotland) (49, 50), and one study in Scotland (51).

In terms of climate change exposure pathways, 18 of the 24 studies assessed health impacts of climate change related hazards, of which 10 reported on increase in ambient temperature (29–31, 34, 35, 38–40, 42, 45), 5 on extreme cold (31, 35, 36, 46, 51), 4 on extreme heat (35, 46, 48, 51), and 4 on heavy rainfall and flooding (32, 33, 37, 47) (4 studies reported on more than one exposure (31, 35, 46, 51)). Only one study assessed health impacts of climate change related health risks (changes to air quality) (49). Five studies assessed health impacts of solutions and responses to address climate change, of which 4 reported on mitigation policies or interventions (41, 43, 50) (one article reported on 2 studies (41)) and one reported on adaptation policies or interventions (44).

In terms of population groups, 20 of the 24 studies reported on people experiencing the greatest deprivation (29–33, 35–37, 39, 41–48, 50, 51), 2 on ethnic minority groups (37, 49), 2 on people experiencing homelessness (38, 40), and one on people with drug and/or alcohol dependence (34) (one study reported on 2 population groups (37)).

A summary of the main findings of each of the included studies can be found in **Table 1**.

**Table 1.**
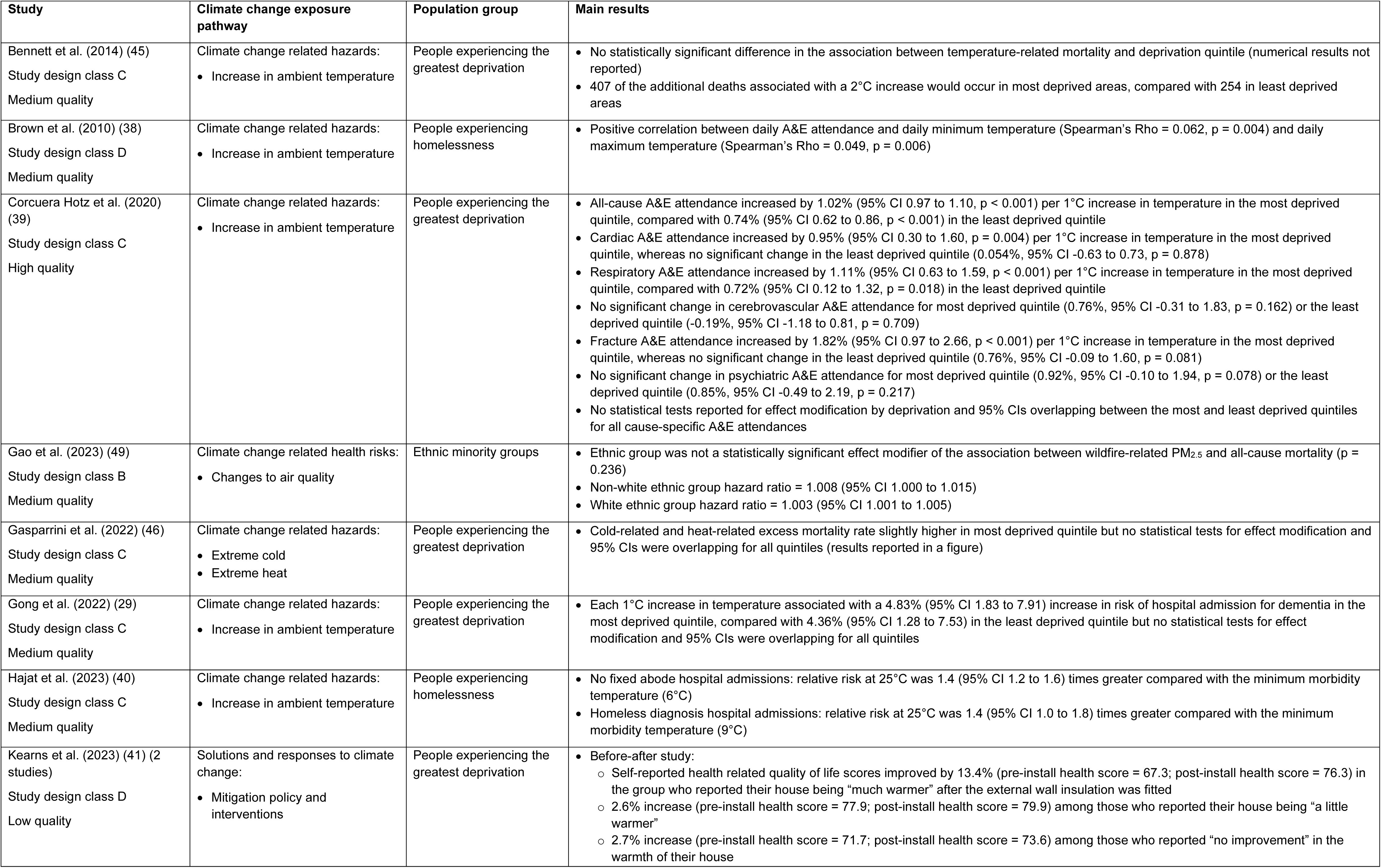
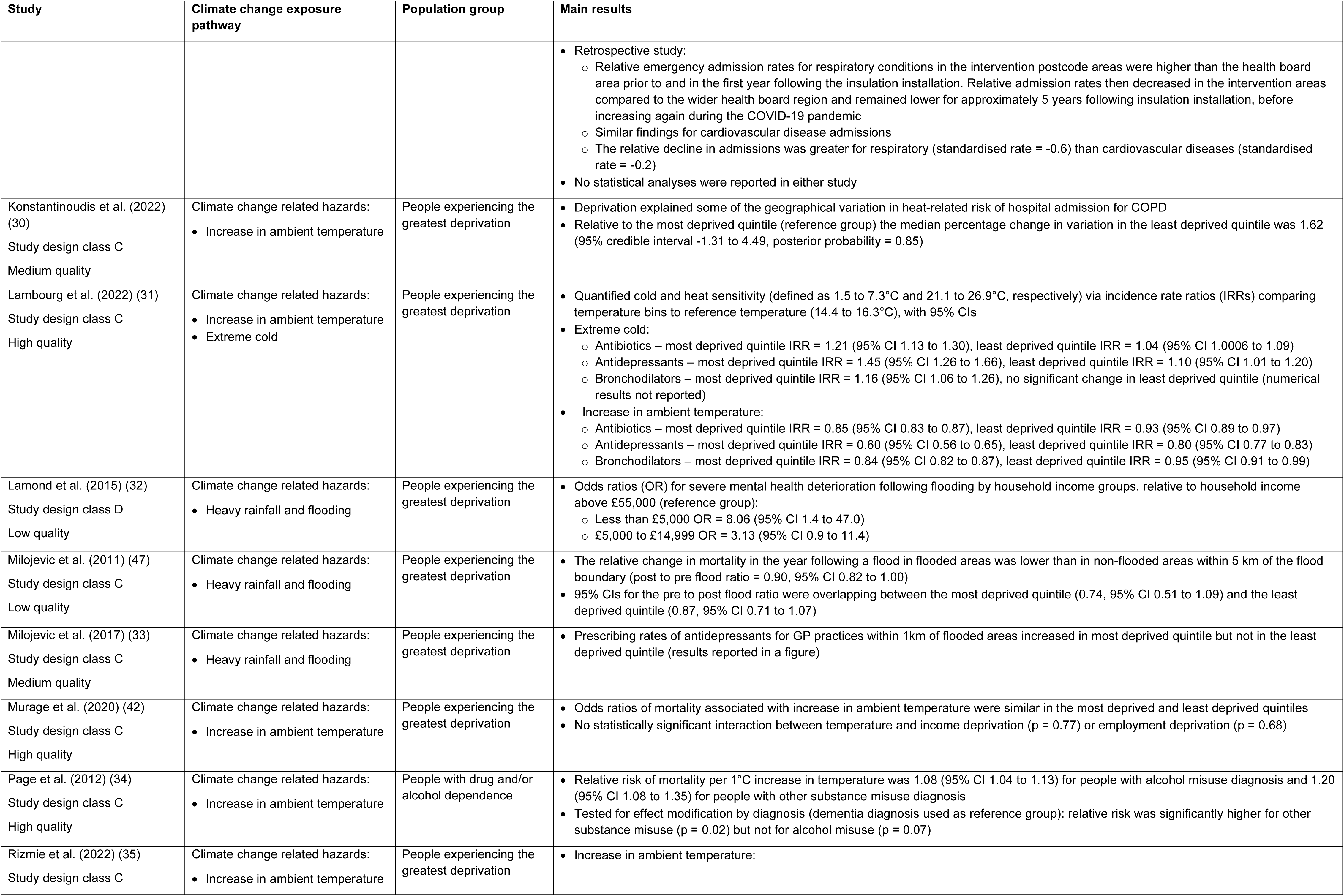
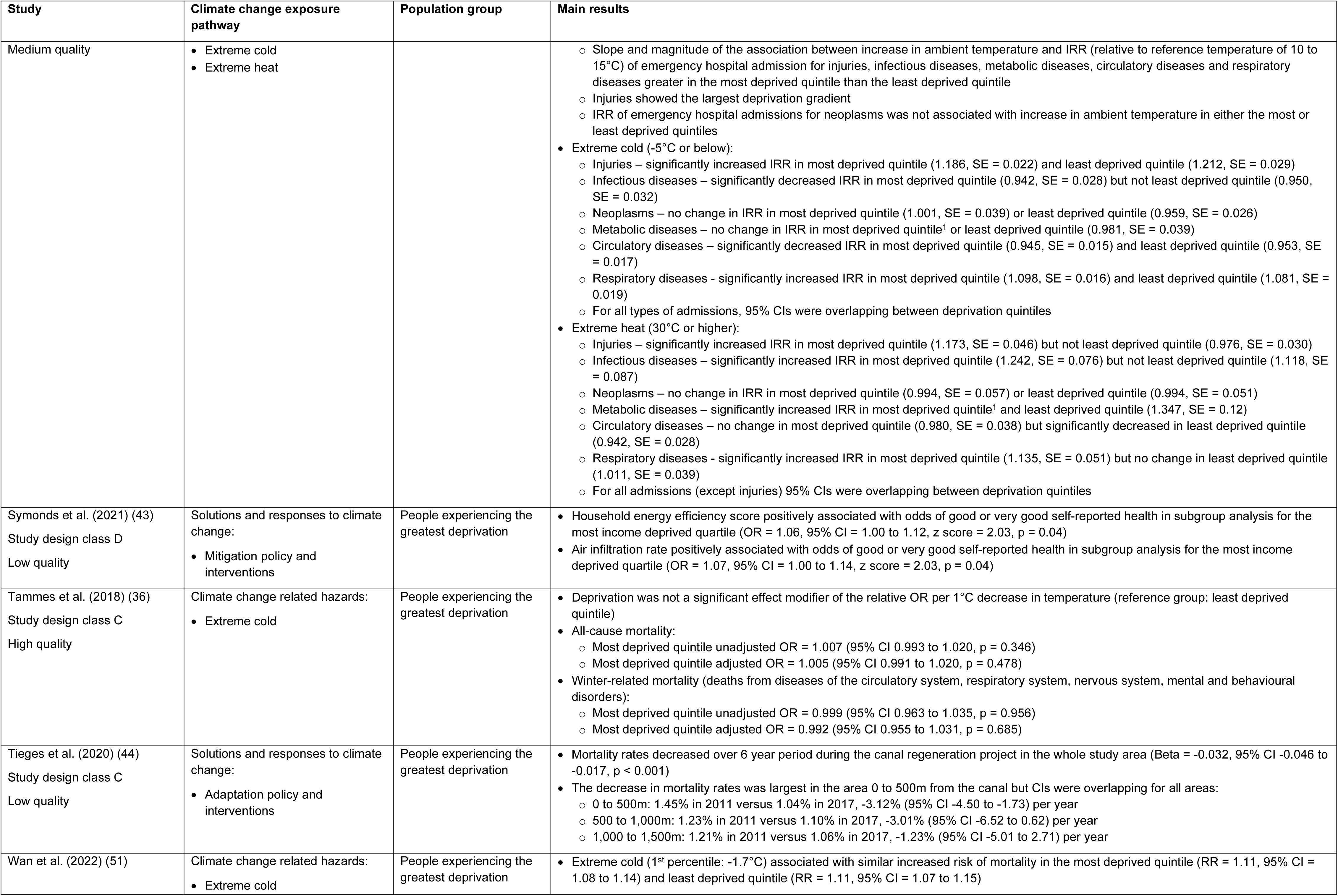
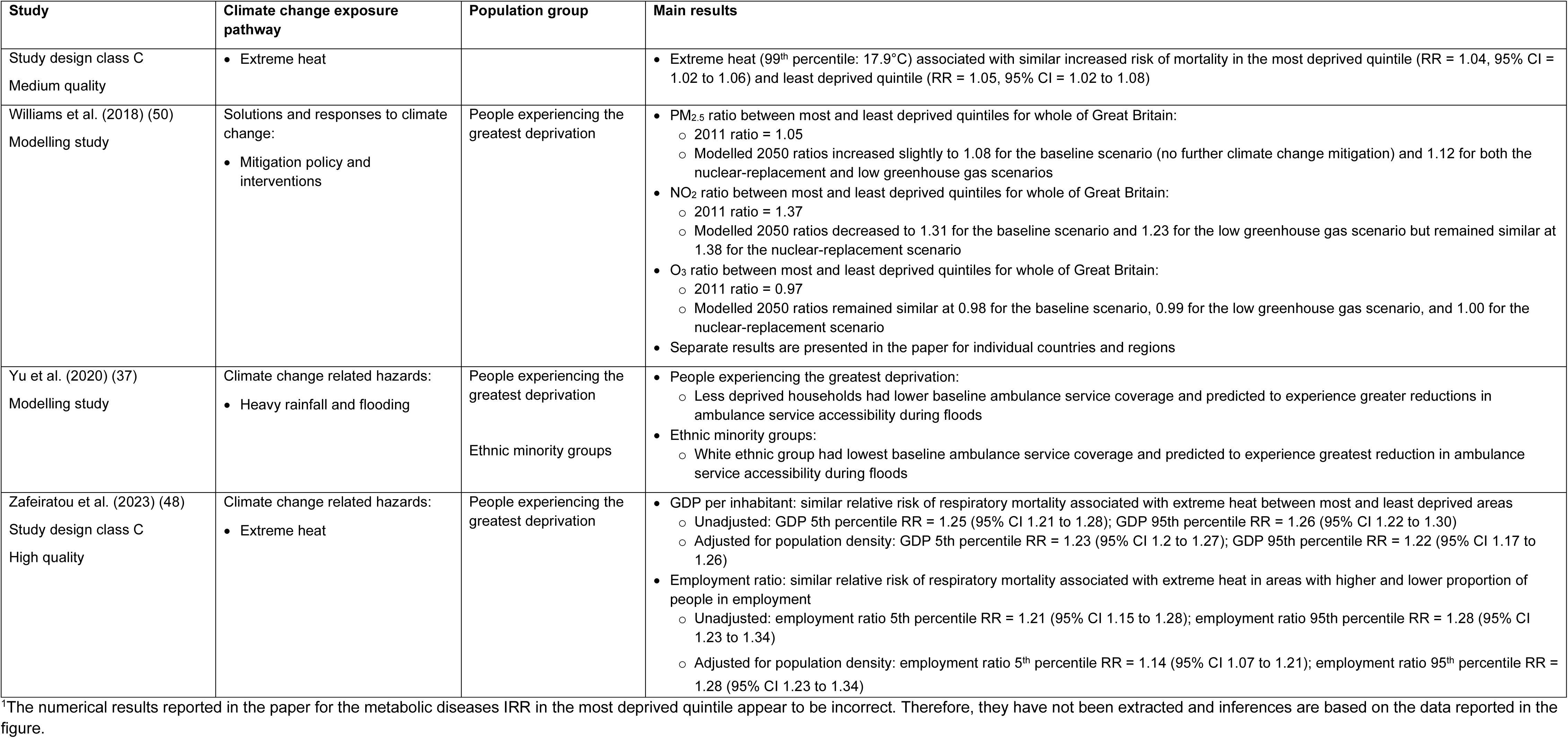
Summary table of the main results of the included studies.

### 3.3. Critical appraisal

Results of the critical appraisal are shown in **Figure 2**. Of the 24 included studies, one was a prospective cohort study (study design class B) which was rated as medium quality (49); 12 were time series studies (study design class C) of which 4 were rated as high quality (31, 34, 39, 48) 6 as medium quality (29, 33, 35, 40, 46, 51) and 2 as low quality (44, 47), 4 were case-crossover studies (study design class C) of which 2 were rated as high quality (36, 42) and 2 as medium quality (30, 45), 3 were retrospective studies (study design class D) one rated as medium quality (38) and 2 as low quality (41, 43), one was a cross-sectional study (study design class D) rated as low quality (32), one was a before-after study (study design class D) rated as low quality (41), and 2 were modelling studies (37, 50) (no study design class or quality rating assigned).

**Figure 2.**
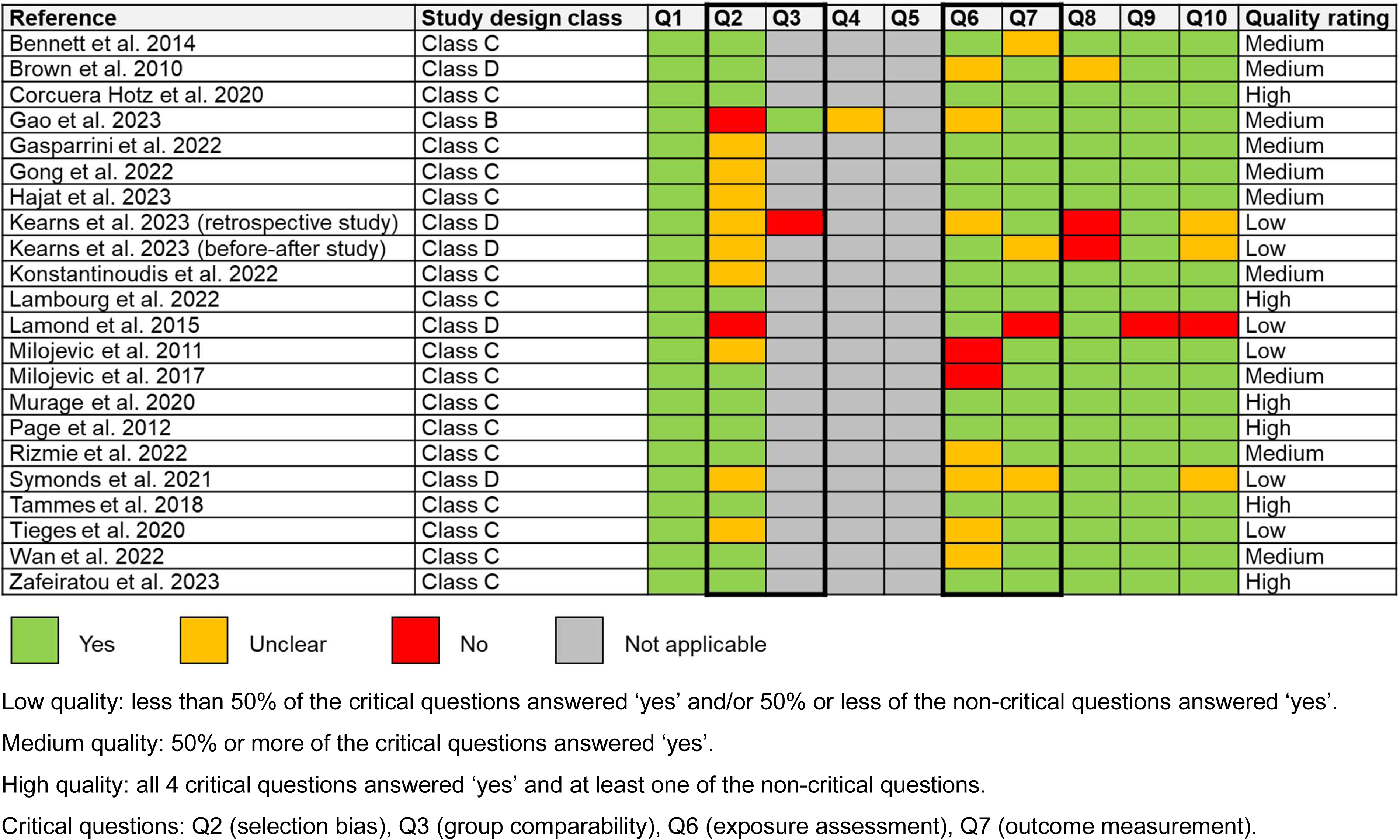
**Study quality assessment of observational studies using the Quality Criteria Checklist**

The study designs of the included studies provided low-level evidence due to high risk of bias associated with the designs used: only one prospective cohort study was identified, and the remaining observational studies were either class C or class D. In addition, the methodological quality of the included studies was mixed, with most studies being rated as medium or low quality. A common limitation identified was the use of area-based measures of deprivation, which may not be an accurate indicator of deprivation levels for people living in those areas. Another frequent limitation was the absence of statistical tests for effect modification, which we have indicated in the narrative summary accordingly as it is therefore unclear whether the results reflect statistically significant differences; in these cases, the narrative summary was based on whether confidence intervals were overlapping.

### 3.4. Evidence gap map

The 24 included studies were mapped onto an interactive evidence gap map available online (16). A screenshot of the map is presented in **Figure 3**.

**Figure 3.**
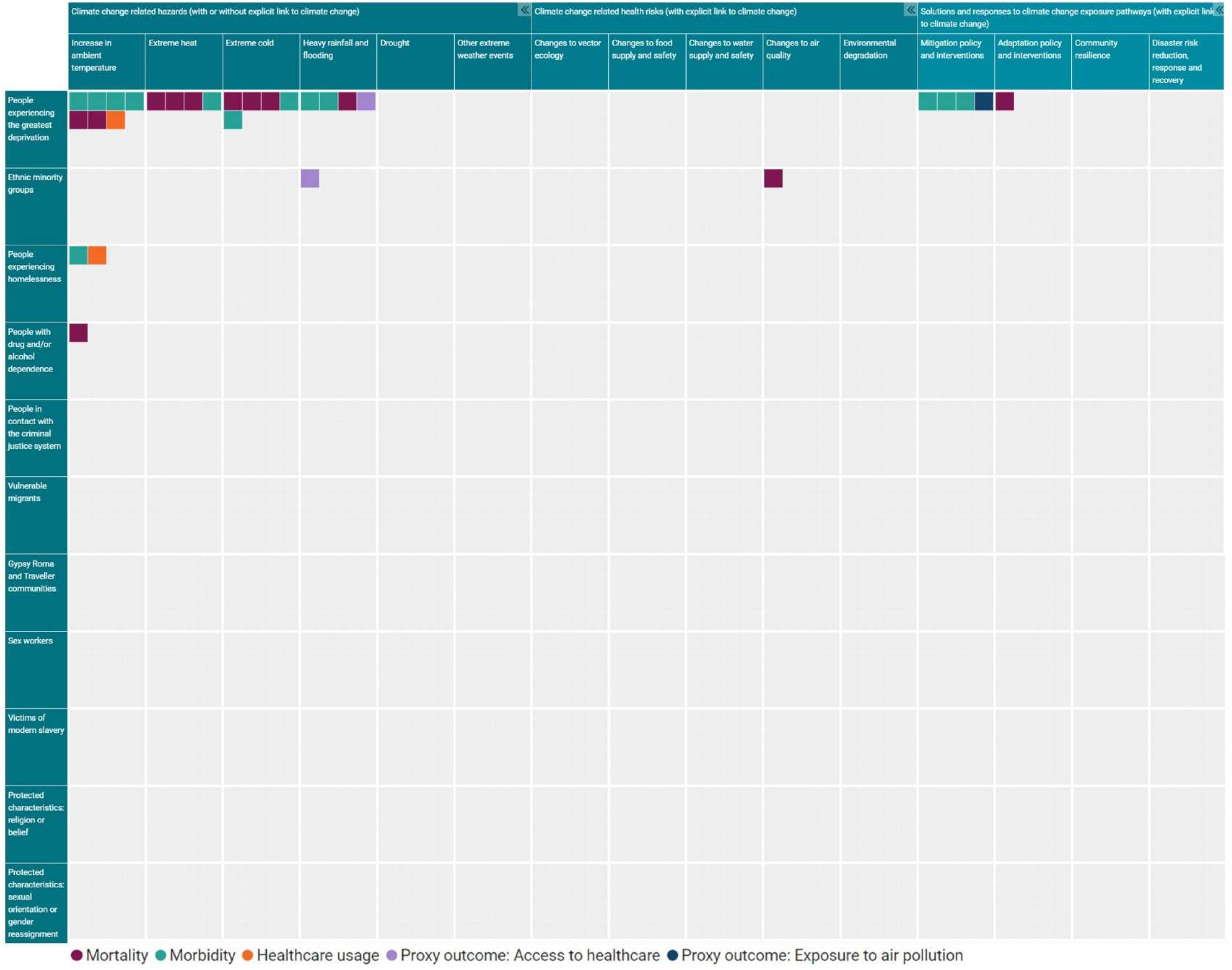
**Screenshot of the interactive evidence gap map**

### 3.5. Evidence on climate change related hazards

Eighteen of the 24 included studies reported on the health impacts of climate change related hazards (see **Table 1** for a summary of the main findings of each study). Of these 18 studies, 10 reported on the health impacts of increase in temperature, 5 of extreme cold, 4 of extreme heat, and 4 of heavy rainfall and flooding. No studies reporting on drought or on other extreme weather events were identified.

In terms of population groups, 15 studies reported on people experiencing the greatest deprivation (29–33, 35–37, 39, 42, 45–48, 51), one of which also reported on ethnic minority groups (37), 2 on people experiencing homelessness (38, 40), and one on people with drug and/or alcohol dependence (34). No studies on climate change related hazards were identified for the other population groups included in this review.

#### 3.5.1. People experiencing the greatest deprivation - increase in ambient temperature

Seven studies reported on the health impacts of increase in ambient temperature for people experiencing the greatest deprivation.

Of these 7 studies, 2 case-crossover studies reported on mortality (42, 45). Findings from both studies suggested that the effect size of the associations between increased temperature and risk of all-cause mortality or cardiorespiratory mortality did not vary depending on the level of area deprivation. However, one of these studies predicted that an increase in temperature would be associated with a larger increase in the number of temperature-related cardiorespiratory deaths in the most deprived areas (45).

Three of the 7 studies (one case-crossover and 2 time series) reported on hospital admissions (29, 30, 35). One of these studies suggested that an increase in ambient temperature may be associated with a greater increase in risk of emergency hospital admissions for some diseases and injuries in the most deprived areas compared to the least deprived areas (35). The second study suggested that area deprivation might modify geographical variation in heat-related hospital admissions for chronic obstructive pulmonary disease (COPD), although the study authors concluded that the evidence of effect was inconclusive (30). The third study found that the increase in hospital admissions for dementia associated with an increase in ambient temperature was similar in the most and least deprived areas (29).

One of the 7 studies was a time series analysis on prescribing rates (31), which found that an increase in ambient temperature was associated with a reduction in prescribing for antibiotics, antidepressants and bronchodilators in both the most and least deprived areas and that the effect size of this reduction may be greater in the most deprived areas compared to the least deprived areas.

The remaining study was a time series analysis on healthcare usage (39), which suggested that an increase in ambient temperature might have a more pronounced relationship with overall (all-cause) A&E attendance in the most deprived areas. Findings for cause-specific A&E attendance were unclear due to greater uncertainty and limited statistical power.

In summary, the evidence was mixed regarding the health equity impacts of increase in ambient temperature for people experiencing the greatest deprivation. The studies on mortality suggested similar impacts for people living in the most deprived and least deprived areas. There was suggestive evidence that the increase in hospital admissions for some diseases and injuries may be larger in more deprived areas. Conversely, one study suggested a greater reduction in prescribing rates for commonly prescribed medications in the most deprived areas. However, all 7 studies were conducted in the general population using an area-level measure of deprivation, of which 4 studies did not perform statistical tests to formally test for deprivation as an effect modifier of the association between increase in ambient temperature and health outcomes (29, 31, 35, 39), which limited the conclusions that could be drawn.

#### 3.5.2. People experiencing the greatest deprivation - extreme cold

Five studies reported on health impacts of extreme cold for people experiencing the greatest deprivation.

Of the 5 studies included, 3 studies (one case-crossover (36) and 2 time series (46, 51)) found that the increased risk of mortality associated with extreme cold was similar for those living in the most and least deprived areas.

One of the 5 studies was a time series analysis on emergency hospital admissions (35), which found that extreme cold was associated with similar changes in the risk of hospital admissions for a range of health conditions in the most and least deprived areas.

The remaining study was a time series analysis (31), which found that extreme cold was associated with increased prescribing rates for antibiotics and antidepressants in both the most and least deprived areas and that the effect size of this increase may be larger in the most deprived areas. Extreme cold was also associated with increased prescriptions for bronchodilators in the most deprived areas but not in the least deprived areas.

In summary, the evidence suggests that the impacts of extreme cold on mortality and hospital admissions were similar for people living in the most deprived and least deprived areas, whereas one study provided some evidence for a greater increase in prescriptions in the most deprived areas. However, all 5 studies were conducted in the general population using an area-level measure of deprivation, of which 4 studies did not perform statistical tests to formally test for deprivation as an effect modifier of the association between extreme cold and health outcomes (31, 35, 46, 51), which limited the conclusions that could be drawn.

#### 3.5.3. People experiencing the greatest deprivation - extreme heat

Four studies reported on the health impacts of extreme heat for people experiencing the greatest deprivation.

Of these 4 studies, 3 were time series analysis of mortality which all found a similar association between extreme heat and all-cause mortality in the most and least deprived areas (46, 48, 51).

The other study was a time series analysis on hospital admissions (35). Findings suggested that extreme heat was associated with similar changes in the risk of emergency hospital admissions for a range of health conditions in the most and least deprived areas. The only exception was injuries, which was the only outcome assessed where confidence intervals were not overlapping between the most and least deprived areas, potentially indicating a larger increase in the most deprived areas.

In summary, the evidence suggests that the association between extreme heat and mortality and hospital admissions for a range of health conditions was similar for people living in the most deprived and least deprived areas, although evidence from one study suggested a greater impact on hospital admissions for injuries in the most deprived areas. However, the conclusions that could be drawn were limited because all 4 studies were conducted in the general population using an area-level measure of deprivation and did not perform statistical tests to formally test for deprivation as an effect modifier of the association between extreme heat and health outcomes.

#### 3.5.4. People experiencing the greatest deprivation - heavy rainfall and flooding

Four studies reported on the health impacts of heavy rainfall and flooding for people experiencing the greatest deprivation.

Of the 4 studies, one time series analysis assessed all-cause mortality (47) and found a similar reduction in mortality in the year following flooding in the most and least deprived areas. The study authors noted that the reduction in mortality was likely due to displacement of people from flooded areas rather than a direct effect of flooding on health.

Two of the 4 studies reported on mental health outcomes. The first study was a time series analysis (33) which found that the prescribing rates for antidepressants were increased in the most deprived areas in the year following floods, whereas there was no change in the least deprived areas. The second study was a cross-sectional study (32), which found that self-reported long-term mental health deterioration among households that had been flooded was more pronounced for those with the lowest household income compared with those with the highest household income.

The remaining study was a modelling study which reported on access to healthcare (37). This study predicted that the least deprived households would experience a greater reduction in ambulance accessibility during flood events than the most deprived households, which the study authors concluded was likely due to a larger proportion of the most deprived households being in urban areas, whereas more of the least deprived households were in rural areas. These results highlight the contribution of geographical factors to inequalities in ambulance accessibility by deprivation.

In summary, there was evidence from 2 studies to suggest greater impacts on mental health among people experiencing the greatest deprivation, although these results should be interpreted with caution as outcomes were self-reported in one study (32) and the other study did not assess whether households had been flooded (33). Conclusions for the other health outcomes were restricted by the limited evidence available: one study suggested similar impacts on mortality by deprivation and one modelling study suggested geographical factors may influence inequalities in ambulance accessibility for people experiencing the least deprivation living in rural areas. Conclusions were further limited by the fact that 2 of the 4 studies were conducted in the general population using area-level measures of deprivation and did not report statistical tests for effect modification by deprivation (33, 47) and only one of the 4 studies assessed whether households had been flooded (32).

#### 3.5.5. Ethnic minority groups - heavy rainfall and flooding

One modelling study assessed the impact of flooding on access to healthcare (37). Results showed a greater predicted reduction in ambulance accessibility during flood events for the white ethnic group than for ethnic minority groups (Asian, Black, Mixed, Other), which the study authors suggested was likely due to more people from ethnic minority groups living in urban areas, whereas more of the white ethnic group lived in rural areas. These findings highlight the role of geographical factors in inequalities in ambulance accessibility by ethnicity, similar to the findings by deprivation reported in section 3.5.4.

In summary, only one study was identified, which suggested that geographical factors may influence inequalities in ambulance accessibility for the white ethnic group living in rural areas.

#### 3.5.6. People experiencing homelessness - increase in ambient temperature

Two studies reported on health impacts of increase in ambient temperature for people experiencing homelessness. Of these, one time series analysis (40) found that increase in ambient temperature was associated with increased risk of all-cause emergency hospital admissions for people experiencing homelessness. The other was a retrospective study (38) which found that increase in ambient temperature was associated with a greater risk of A&E attendance for people experiencing homelessness.

In summary, evidence from 2 studies suggested that increase in ambient temperature increased risk of adverse health outcomes (hospital admission and A&E attendance) for people experiencing homelessness. However, conclusions could not be drawn regarding the disproportionate impact on health for this group due to the lack of comparison to a control group of people not experiencing homelessness.

#### 3.5.7. People with drug and/or alcohol dependence - increase in ambient temperature

One time series analysis reported on the association between increase in ambient temperature and all-cause mortality for people with alcohol misuse and people with other substance misuse (34). Results suggested that there was a positive association between increased ambient temperature and risk of mortality among people with alcohol misuse and people with other substance misuse.

In summary, only one study was identified, which found an increased risk of mortality for people with drug and/or alcohol dependence with increased temperature, but as no comparison was made to a control group of people without drug and/or alcohol dependence, it was not possible to draw conclusions about whether increase in ambient temperature disproportionately affects health for this population group.

### 3.6. Evidence on climate change related health risks

Only one study reported on the health impacts of climate change related health risks. This study assessed health impacts of changes to air quality for ethnic minority groups (49) (see **Table 1** for a summary of the main findings).

No studies were identified for the other climate change related health risks (changes to vector ecology, changes to food supply and safety, changes to water supply and safety, or environmental degradation) or the other population groups included in this review.

#### 3.6.1. Ethnic minority groups - changes to air quality

One prospective cohort study assessed the association between wildfire-related PM_2.5_ levels and all-cause mortality. Results showed that ethnicity (white ethnicity versus non-white ethnicity) was not a statistically significant effect modifier of this association (49). However, there was more uncertainty for the non-white group due to smaller sample size and the study authors did not report power calculations to determine whether the study was sufficiently powered to detect an interaction.

In summary, only one study was identified, and it was not possible to draw conclusions about the health equity impacts of climate change related changes to air quality for ethnic minority groups due to the limited evidence available and a potential lack of statistical power in this study.

### 3.7. Evidence on solutions and responses to address climate change

Five studies reported on health impacts of solutions and responses to address climate change. All 5 studies reported on people experiencing the greatest deprivation. Of these 5 studies, 4 reported on mitigation policy and interventions (41, 43, 50) (one article reported on 2 studies (41)) and one on adaptation policy and interventions (44) (see **Table 1** for a summary of the main findings of each study).

No studies were identified for the other solutions and responses to address climate change (community resilience or disaster risk reduction, response, and recovery) or the other population groups included in this review.

#### 3.7.1. People experiencing the greatest deprivation - mitigation policy and interventions

Four studies reported on the health impacts of climate change mitigation policy and interventions for people experiencing the greatest deprivation. Of these, 3 studies reported on the association between household energy efficiency and health outcomes (41, 43) (one article reported on 2 studies (41)). One of these studies was a before-after study (41), which found that an intervention to improve home energy efficiency in a deprived area may be associated with improved self-reported health-related quality of life among participants who reported their home being warmer following the intervention. However, the lack of a control group limits the extent to which this finding can be attributed to the intervention. In addition, as the intervention was only conducted in deprived areas, there was no comparison with the impact on health in less deprived areas. Another retrospective study (reported in the same paper) suggested a greater reduction in the rate of hospital admissions for cardiovascular and respiratory conditions in the intervention area compared with the remainder of the health board area. However, both the intervention area and comparison area had high levels of deprivation: respectively, 73% and 54% of areas were in the lowest 2 quintiles of the Scottish IMD. In both studies, it is unclear whether these changes were statistically significant due to a lack of statistical analyses. The third study was a retrospective study (43), which found that better home energy efficiency and air infiltration rates were both positively associated with the proportion of people reporting good or very good health in the most deprived areas. However, the study did not compare the effect size of this association with less deprived areas.

The other study was a modelling study (50), which found that the impact of climate change mitigation scenarios on inequalities in exposure to air pollution for people living in deprived areas differed depending on the scenario and type of pollutant. A scenario using nuclear power to replace fossil fuels was predicted to increase inequalities in exposure to PM_2.5_ by 2050 compared with 2011, whereas no change in inequalities was predicted for exposure to NO_2_ or O_3_. A low greenhouse gas scenario was also predicted to increase inequalities in exposure to PM_2.5_ but to decrease inequalities in exposure to NO_2_, with no change for O_3_. However, the study also predicted that a baseline scenario (no further climate change mitigation measures) would lead to increased inequalities in exposure to PM_2.5_, decreased inequalities in exposure to NO_2_ and no change in inequalities for O_3_.

In summary, evidence from 3 studies reporting on climate change mitigation policies that improve home energy efficiency suggested that there may be health co-benefits for people experiencing the greatest deprivation. However, these results should be interpreted with caution because of important methodological limitations, including the potential for self-report bias and ecological bias because deprivation was measured at the area-level. In addition, no conclusions could be drawn about the impact of these interventions on health equity due to the lack of comparison with less deprived areas. Only one study (modelling) was identified on climate change policies aimed at reducing emissions and no conclusions could be drawn due to the limited evidence available.

#### 3.7.2. People experiencing the greatest deprivation - adaptation policy and interventions

Only one study reported on the health impacts of climate change adaptation policy and interventions for people experiencing the greatest deprivation (44). This time series analysis investigated the health benefits of a canal regeneration project aimed to improve climate change adaptation in an area with high levels of deprivation. Results suggested a greater reduction in all-cause mortality in areas nearest to the canal compared with areas further away from the canal, but there was no comparison between the impacts on health between the most and least deprived areas. In addition, it is unclear whether this was statistically significant due to a lack of statistical tests. Moreover, the extent to which this reduction can be attributed to the intervention is unclear given a general reduction in mortality over time in the study area.

In summary, only one study was identified, and it was not possible to draw conclusions about the impact on people experiencing the greatest deprivation due to the limited evidence available, methodological limitations, and the lack of comparison with a less deprived area.

## 4. Discussion

### 4.1. Main findings

This rapid systematic review included 24 studies examining the health equity impacts of climate change in the UK. The conclusions that could be drawn from the existing evidence were limited by the small number of studies that assessed the same exposure-outcome association for each population group. Most studies provided low-level evidence in terms of study design and had important methodological limitations.

Twenty of the 24 studies included in this review reported on people experiencing the greatest deprivation. The findings tended to suggest that the impacts of increase in ambient temperature, extreme heat, and extreme cold on mortality were similar for people living in the most deprived and least deprived areas. The impacts on hospital admissions were more heterogenous, suggesting a potential greater impact of increase in temperature for some health conditions for those living in the most deprived areas but no difference by deprivation for extreme cold or for extreme heat (with the exception of hospital admissions for injuries in extreme heat). There was also suggestive evidence from 3 studies for potential health co-benefits of climate change mitigation interventions for people experiencing the greatest deprivation, although this was based on low-level evidence and the impact on health equity could not be assessed as there were no comparisons with less deprived groups. Conclusions for the remaining outcomes and exposures could not be drawn due to the limited evidence identified in terms of number of studies and study designs.

Overall, the conclusions that could be drawn for people experiencing the greatest deprivation were limited. Despite not limiting our definition of deprivation to IMD (for instance, terms related to income and unemployment were included in our search strategy, see Supplementary Material 4), 15 of the 20 studies were conducted in the general population using area-based measures of deprivation to perform subgroup analysis, with a further 3 studies conducted in an area experiencing high levels of deprivation. Therefore, the findings of these studies cannot be generalised to individuals within those areas because area-based measures may not provide an accurate representation of deprivation at the individual level (52, 53). In addition, most studies did not perform statistical tests to formally test for deprivation as an effect modifier, or did not compare with less deprived areas, which precluded conclusions being drawn on the disproportionate impact by deprivation.

Two studies assessing the impact of increase in ambient temperature on people experiencing homelessness were identified, suggesting an increased risk of hospital admission and A&E attendance, although conclusions could not be drawn regarding the disproportionate impact on health for this group due to the lack of a control group of people not experiencing homelessness.

No conclusions could be drawn for ethnic minority groups (one study on heavy rainfall and flooding and one on changes to air quality), or people with drug and/or alcohol dependence (one study on increase in ambient temperature) due to the limited evidence identified, both in terms of number of studies and methodological quality.

No studies were identified for the other population groups of interest for this review, indicating that the lack of review-level evidence identified in our initial scoping exercise (17) reflects a paucity of primary research. In addition, no studies were identified that were conducted in settings associated with Core20PLUS population groups (prisons and places of detention, asylum seeker accommodation settings, traveller sites, temporary housing accommodation, homeless shelters, and rehabilitation centres) or that assessed the mediating pathways underlying increased vulnerability to the health impacts of climate change exposure pathways. This highlights an important evidence gap for future research.

In terms of the health outcomes assessed, most of the evidence identified reported on morbidity (10 studies) or mortality (10 studies), such as cardiovascular and respiratory outcomes. Other outcomes that were assessed included mental health (3 studies), healthcare usage (2 studies on A&E attendance), and proximal determinants of health (2 studies: one on access to healthcare and one on exposure to air pollution).

### 4.2. Implications of this study

This rapid systematic review adds to the body of literature on the health equity impacts of climate change in the UK by identifying several evidence gaps in the existing evidence base. Researchers should consider these gaps and, when possible, design studies that address the methodological limitations of the existing evidence identified in this review, such as using prospective cohort study designs, conducting statistical tests for effect modification, and including individual-level measures of deprivation and exposure assessment. Indeed, the Health Effects of Climate Change (HECC) report recommends that health equity should be embedded in all research into the health impacts of climate change (2). This will be essential to identify those most vulnerable to the health impacts of climate change as well as considering potential health co-benefits and unintended negative consequences of climate change adaptation and mitigation interventions on vulnerable groups to inform and target future action appropriately.

### 4.3. Strengths and limitations

To our knowledge, this is the first review to perform a comprehensive and systematic assessment and summary of the available evidence on the health equity impacts of climate change in the UK, encompassing 15 climate change exposure pathways and 12 Core20PLUS groups (including associated settings) to summarise the available evidence and highlight evidence gaps. By using the Core20PLUS framework (14) to define the population groups eligible for inclusion in this review, the findings can be used to inform efforts to improve health equity in health security in the UK (15) by identifying population groups at-risk of health inequalities associated with climate change and groups for whom further research is needed.

Another strength is that extensive searches were undertaken to identify relevant evidence (5 databases and 11 grey literature sources), including health equity focused sources such as the FAIR database and Climate Just. In total, more than 20,000 records were screened, demonstrating the breadth of literature considered for inclusion.

A limitation of this rapid systematic review is the use of streamlined methodologies to accelerate the review process. For example, rather than being done independently by 2 reviewers, some stages (such as full text screening and data extraction) were done by one reviewer and checked by another. To mitigate the risk of potential errors arising from not performing all stages in duplicate, extensive quality assurance was conducted. Another limitation is that critical appraisal was conducted at the study level to provide an indication of methodological quality; we did not assess the risk of bias for each outcome.

Finally, given the wide scope of this review, studies addressing climate change related health risks or solutions and responses were required to explicitly establish a connection to climate change to be included. This was necessary to maintain the focus on climate change because there is an extensive literature on the health impacts of air pollution and interventions to improve air quality that are not due to climate change, which was beyond the scope of this review.

### 4.2 Conclusions

Most of the available evidence assessed the health equity impacts of climate change related hazards by deprivation, suggesting that impacts of increase in ambient temperature, extreme heat, and extreme cold on mortality may be similar for people living in the most deprived and least deprived areas. However, only limited conclusions could be drawn for other outcomes and exposures due to methodological limitations (such as area-level measures of deprivation) and heterogeneity in the health outcomes assessed in relation to people experiencing the greatest deprivation.

There was also some evidence suggesting that increase in ambient temperature was associated with an increased risk of hospital admission and A&E attendance for people experiencing homelessness, although conclusions could not be drawn regarding the disproportionate impact on health for this group due to the lack of a control group.

This rapid systematic review of the available evidence on the health equity impacts of climate change in the UK identified several evidence gaps and limitations in the evidence base for the other Core20PLUS population groups and climate change exposure pathways included in this review.

This highlights the need for future research to address these evidence gaps and to consider the methodological limitations identified in this review, including using prospective cohort study designs, conducting statistical tests for effect modification, and including individual-level measures of deprivation and exposure assessment.

## Funding

This study did not receive any specific grant from funding agencies in the public, commercial or not-for-profit sectors.

## Data availability statement

All data that support the findings of this study are included within the article, its supplementary files, and the original mapping review report (https://www.gov.uk/government/publications/health-equity-impacts-of-climate-change).

## Author contributions

**Matthew Bosworth:** Methodology, Investigation, Visualisation, Project administration, Writing – Original Draft. **Zalaya Simmons:** Conceptualisation, Methodology, Investigation, Visualisation, Project administration. **Rhiannon Cordiner:** Investigation, Writing – Review & Editing. **Tabitha Kavoi:** Investigation. **Beti Evans:** Investigation. **Tara Quinn:** Investigation, Writing – Review & Editing. **Barbara Rubiell:** Writing – Review & Editing. **Amy Jackson:** Conceptualisation, Writing – Review & Editing. **Ahimza Nagasivam:** Conceptualisation. **Nicola Pearce-Smith:** Methodology, Investigation, Writing – Review & Editing. **Lea Berrang-Ford:** Writing – Review & Editing. **Daphne Duval:** Conceptualisation, Methodology, Supervision, Writing – Original Draft.

## Acknowledgements

We would like to thank colleagues from UKHSA in the Research, Evidence and Knowledge division (especially Tom Pink, Angela Page, Dorothy Hirsch, Carolina Arevalo, and Wendy Marsh), the Centre for Climate Change and Health Security (especially Helen Macintyre, Jennifer Israelsson, and Andrew Fox), and Jennifer Hill from the All Hazards Public Health Response division for their input into specific aspects of this review.

## Conflict of interest

The authors declare that they have no competing interests.

## Disclaimer

The views expressed in this article are those of the authors and are not necessarily those of the UK Health Security Agency or the Department of Health and Social Care.

## Notes

### Competing Interest Statement

The authors have declared no competing interest.

