## Supplementary materials for "Health equity impacts of climate change in the UK: a rapid systematic review and interactive evidence gap map"

### Supplementary Material 1. Modifications to the protocol

#### Inclusion and exclusion criteria

In the rapid mapping review protocol, 'social housing accommodation' and 'council housing estates' were listed as eligible for inclusion as these settings were thought to be relevant for identifying people experiencing the greatest deprivation. However, these settings were removed from the inclusion criteria at the screening stage as they were not reliable indicators of people experiencing the greatest deprivation due to the socio-economic variability among residents (1).

Our protocol aimed to include evidence for people experiencing the greatest deprivation, based on the most deprived 20% of the population defined by the IMD (in line with the Core20PLUS framework), as well as other measures of deprivation. Once the screening started, we agreed to also include studies that measured deprivation based on quartiles of deprivation (most deprived 25% of the population) to ensure that no relevant evidence on the most deprived populations was excluded.

Two additional proxy outcomes were added to the inclusion criteria for risky health behaviours and physical activity. These were included to capture a broader spectrum of determinants that may influence health outcomes related to climate change.

Other minor refinements were made to the eligible outcomes based on insights gained during the screening process. To provide clarity on the specific mental health outcomes that were eligible for inclusion, some examples of eligible outcomes were added to the protocol: "(including, but not limited to, self-reported or clinical measures of stress, anxiety, depression, obsessive compulsive disorder, phobias, psychological distress, post-traumatic stress disorder, eating disorders, substance abuse disorders, personality disorders, resilience, and quality of life)". Happiness and life satisfaction were also added as examples for the excluded outcome of well-being. This distinction was made to illustrate the difference between mental health outcomes and broader measures of well-being.

#### Sources searched

The FAIR database and the King's Fund Library were not included in the databases listed in the rapid mapping review protocol. These were added after the protocol was published on the recommendation of 2 information scientists.

We excluded 4 of the grey literature sources listed in the protocol due to their limited evidence in relation to the review question because:

- the Health Inequalities Portal and Institute of Health Equity lacked evidence on climate change
- the Adaptation Clearinghouse focused on US-contexts
- the Collaboration for Environmental Evidence Database provided mostly review-level evidence which was out of scope for this review

#### Data extraction

The rapid mapping review protocol stated that we would extract data on the mediating pathways underlying the health equity impacts of climate change and, depending on the evidence identified, map the corresponding findings. However, due to the lack of evidence identified (no qualitative studies identified and none of the quantitative studies conducted formal mediation analysis), we decided not to extract data on mediating pathways for this mapping review.

#### Critical appraisal

The use of study design class to provide an indication of the potential for bias within each study design was not planned in the rapid mapping review protocol. This was done in order to add clarity and support discussion around the level of evidence. Critical appraisal was done independently in duplicate by 2 reviewers instead of being done by one reviewer and checked by a second as planned in the protocol.

#### Synthesis

In the rapid mapping review protocol, it was suggested that, depending on the type of evidence identified, the third dimension of the evidence gap map would be either the mediating pathways or study quality. Following discussion within the review team, including topic experts, we decided to map the health outcomes in the third dimension as this was deemed to be more informative.

The protocol also stated that 2 maps would be produced: one for climate change related hazards and health risks, and another for solutions and responses to climate change. As the number of studies identified was low (only 24 studies), we decided to create one map containing all of the exposure pathways.

Supplementary Material 2. Inclusion and exclusion criteria

| | Included | Excluded |
|---|---|---|
| **Country** | UK | Non-UK |
| **Population** | Core20PLUS population groups included within this review:<br>• people experiencing the greatest deprivation (the most deprived 20% as defined by IMD, also other deprivation measures will be considered)<br>• ethnic minority groups (protected characteristics)<br>• people from protected characteristics groups related to gender reassignment and sexual orientation (including LGBTQ+: lesbian, gay, bisexual, transgender, intersex, queer or questioning, asexual and many other terms such as non-binary and pansexual)<br>• people from protected characteristics groups related to religion or belief<br>• people experiencing homelessness<br>• people with drug and/or alcohol dependence<br>• people in contact with the criminal justice system<br>• vulnerable migrants<br>• Gypsy Roma and Traveller communities<br>• sex workers<br>• victims of modern slavery<br>• other groups with experience of social exclusion<br>Studies conducted in the general population, but which included a focus on sub-group populations | Core20PLUS population groups not included within this review:<br>• populations with other protected characteristics including age, disability, pregnancy and maternity, and sex<br>• populations with pre-existing health conditions<br><br>Place-based vulnerability, including:<br>• rural versus urban<br>• coastal settings<br>• regional differences |

| | | |
|---|---|---|
| | corresponding to the groups of interest were considered for inclusion | |
| **Settings** | All settings with a focus on settings specific to the population groups of interest, including (but not limited to):<br>• prisons and places of detention including adult prisons, the children and young people's secure estate, approved premises and immigration removal centres<br>• asylum seeker accommodation settings (including, arrival centres, bridging hotels)<br>• temporary housing accommodation<br>• homeless shelters and hostels<br>• rehabilitation centres, drug and alcohol treatment facilities<br>• traveller sites (including caravan sites) | |
| **Intervention or exposure** | Climate change induced events and exposure pathways, including:<br>• climate change related hazards (with or without explicit link to climate change)<br> ○ increase in ambient temperature<br> ○ extreme heat<br> ○ extreme cold<br> ○ heavy rainfall and flooding<br> ○ drought<br> ○ other extreme weather events (such as storms or wildfires)<br>• climate change related health risks with explicit link to climate change | Climate change related health risks and solutions or response without an explicit mention or link to climate change (for instance, studies reporting on changes to vector ecology without a specific link or mention to climate change) were excluded |

| | | |
|---|---|---|
| | ○ changes to vector ecology<br>○ changes to food supply and safety<br>○ changes to water supply and safety<br>○ changes to air quality (including air pollution and aeroallergens and due to emissions ozone or particulate concentration)<br>○ environmental degradation<br>• solutions and responses to climate change exposure pathways with explicit link to climate change<br>○ mitigation policy and interventions<br>○ adaptation policy and interventions<br>○ community resilience<br>○ disaster risk reduction, response and recovery | |
| **Outcomes** | • Observed and projected health effects associated with climate change exposure pathways, including (but not limited to):<br>○ mortality (all-cause and cause-specific)<br>○ respiratory disease<br>○ cardiovascular diseases<br>○ other chronic diseases<br>○ maternal and child outcomes<br>○ mental health (including but not limited to self-reported or clinical measures of stress, anxiety, depression, obsessive compulsive disorder, phobias, psychological distress, post-traumatic stress disorder, eating disorders, substance abuse disorders, personality disorders, resilience, quality of life) | • Non-health related outcomes, such as economic outcomes<br>• Studies focusing on climate change without health outcomes<br>• Wellbeing outcomes (happiness and life satisfaction and so on)<br>• Non-human health outcomes<br>• Socio-economic determinants of health |

| | | |
|---|---|---|
| | - other morbidity outcomes<br>- healthcare usage such as A&E visits or 999 calls<br>- projected health measures such as health impact scores or life-years gained<br>• The following proxy outcomes (proximal determinants of health) were considered for inclusion:<br>- exposure to air pollution<br>- unsafe working conditions where there is a clear link made to at least one direct health risk<br>- poor nutrition<br>- exposure to infectious diseases and vectors<br>- exposure to poor water quality<br>- poor sanitation<br>- access to healthcare<br>- physical activity<br>- risky health behaviours (such as smoking, alcohol and drugs consumption) | |
| **Language** | English | Non-English studies |
| **Date of publication** | 1 January 2010 to February 2024 | Studies published before January 2010 |
| **Study design** | • Observational studies (cohort, case-control, cross-sectional studies and surveillance studies)<br>• Case reports and case series<br>• Ecological studies<br>• Mixed-method studies<br>• Qualitative studies | • Systematic or narrative reviews<br>• Guidelines<br>• Opinion pieces<br>• Modelling studies that use hypothetical data, or non-UK data, to project the health effect of climate change exposure pathways |

| | | |
|---|---|---|
| | • Modelling studies that used UK data to project the health effects of climate change exposure pathways on populations in the UK | |
| **Publication type** | • Peer-reviewed<br>• Preprint<br>• Grey literature (including reports published by government agencies, local government, and non-governmental organisations) | • Conference abstracts<br>• News articles |

Supplementary Material 3. List of grey literature sources searched

Climate Change and Human Health Literature Portal

Health Observatory Resource Catalogue

FAIR database

King’s Fund Library

National Grey Literature Collection

Adaptation Scotland

Climate and Health (Wellcome)

Climate Change Just (Joseph Rowntree Foundation)

Climate Change Committee

COP26 university network

Environment and climate crisis (NESTA)

Supplementary Material 4. Database search strategies

**Search strategy for Ovid Medline ALL**

| | |
|---|---|
| 1 | (deprivation or deprived or disadvantage* or vulnerable).tw,kf. |
| 2 | social risk*.tw,kf. |
| 3 | (sociodemographic* or socioeconomic*).tw,kf. |
| 4 | (inclusion health group* or socially excluded).tw,kf. |
| 5 | working class*.tw,kf. |
| 6 | poverty.tw,kf. |
| 7 | house* income*.tw,kf. |
| 8 | unemployed.tw,kf. |
| 9 | ethnic minorit*.tw,kf. |
| 10 | (Asian British or Black British or South Asian* or Afr* Caribbean*).tw,kf. |
| 11 | protected characteristic*.tw,kf. |
| 12 | (religion or belief*).tw,kf. |
| 13 | sexual orientation.tw,kf. |
| 14 | LGBTQ+.tw,kf. |
| 15 | (lesbian or gay).tw,kf. |
| 16 | (bisexual or intersex or queer or asexual or CIS or non-binary or pansexual).tw,kf. |
| 17 | gender diverse.tw,kf. |
| 18 | transgender.tw,kf. |
| 19 | (homeless* or rough sleep* or vagrant*).tw,kf. |
| 20 | (substance* adj2 (use* or abuse* or misuse or addict* or depend*)).tw,kf. |
| 21 | (drug* adj2 (use* or abuse* or misuse or addict* or depend*)).tw,kf. |
| 22 | (alcohol* adj2 (use* or abuse* or misuse or addict* or depend*)).tw,kf. |
| 23 | (alcoholic* or alcoholism).tw,kf. |
| 24 | (migrant* or immigrant*).tw,kf. |
| 25 | (gypsy or gypsies).tw,kf. |
| 26 | Roma.tw,kf. |
| 27 | (travelling communit* or traveller* or traveler*).tw,kf. |
| 28 | (sex work* or prostitut*).tw,kf. |
| 29 | justice system*.tw,kf. |
| 30 | modern slave*.tw,kf. |
| 31 | (forced labour or forced labor).tw,kf. |
| 32 | human trafficking.tw,kf. |
| 33 | enslavement.tw,kf. |
| 34 | (prisoner* or offend* or remand* or incarcarat* or imprisonment or custod*).tw,kf. |
| 35 | asylum seeker*.tw,kf. |
| 36 | refugee*.tw,kf. |
| 37 | displaced person*.tw,kf. |
| 38 | social deprivation/ |
| 39 | Low Socioeconomic Status/ |
| 40 | Poverty/ |
| 41 | "Ethnic and Racial Minorities"/ |
| 42 | exp Socioeconomic Factors/ |
| 43 | Religion/ |
| 44 | Minority Groups/ |
| 45 | Culture/ |
| 46 | exp "Sexual and Gender Minorities"/ |

| | |
|---|---|
| 47 | Sex Characteristics/ |
| 48 | exp Homosexuality/ |
| 49 | Ill-Housed Persons/ |
| 50 | Substance-Related Disorders/ |
| 51 | Alcohol-Related Disorders/ |
| 52 | Drug Users/ |
| 53 | Alcoholics/ |
| 54 | Vulnerable Populations/ |
| 55 | "Transients and Migrants"/ |
| 56 | Roma/ |
| 57 | Sex Workers/ or Sex Work/ |
| 58 | Prisoners/ |
| 59 | exp Crime Victims/ |
| 60 | exp Criminals/ |
| 61 | "Emigrants and Immigrants"/ |
| 62 | Enslaved Persons/ |
| 63 | Refugees/ |
| 64 | Working Poor/ |
| 65 | Enslavement/ |
| 66 | Human Trafficking/ |
| 67 | or/1-66 |
| 68 | (prison or prisons or jail* or gaol* or detention or correctional).tw,kf. |
| 69 | ((camp* or tent or tents) adj2 (transit* or temporary or pitch* or site*)).tw,kf. |
| 70 | campsite*.tw,kf. |
| 71 | ((asylum or migrant* or refugee* or immigrant* or immigration) adj2 (centre* or center* or camp* or support* or removal*)).tw,kf. |
| 72 | caravan*.tw,kf. |
| 73 | (park home* or mobile home*).tw,kf. |
| 74 | trailer park*.tw,kf. |
| 75 | outreach program*.tw,kf. |
| 76 | shelter*.tw,kf. |
| 77 | (temporary hous* or "no fixed abode").tw,kf. |
| 78 | temporary accommodation.tw,kf. |
| 79 | (supported hous* or social hous* or half-way house* or council hous*).tw,kf. |
| 80 | hostel*.tw,kf. |
| 81 | (food bank* or foodbank*).tw,kf. |
| 82 | soup kitchen*.tw,kf. |
| 83 | (rehabilitation centre* or rehabilitation centre*).tw,kf. |
| 84 | ((substance or drug* or alcohol*) and treatment clinic*).tw,kf. |
| 85 | (detoxification centre* or detoxification center*).tw,kf. |
| 86 | (drug* counsel* or alcohol* counsel*).tw,kf. |
| 87 | harm reduction program*.tw,kf. |
| 88 | sober living home*.tw,kf. |
| 89 | needle exchange program*.tw,kf. |
| 90 | methadone clinic*.tw,kf. |
| 91 | (GUM or genitourinary medicine clinic*).tw,kf. |
| 92 | (red light district or brothel*).tw,kf. |
| 93 | sexual health clinic*.tw,kf. |
| 94 | Refugee Camps/ |

| | |
|---|---|
| 95 | Emergency Shelter/ |
| 96 | Prisons/ |
| 97 | Correctional Facilities/ |
| 98 | Public Housing/ |
| 99 | Rehabilitation Centers/ |
| 100 | Substance Abuse Treatment Centers/ |
| 101 | Counseling/ |
| 102 | Harm Reduction/ |
| 103 | Needle-Exchange Programs/ |
| 104 | or/68-103 |
| 105 | 67 or 104 |
| 106 | climat*.tw,kf. |
| 107 | extreme heat.tw,kf. |
| 108 | (temperature adj increase*).tw,kf. |
| 109 | ambient temperature*.tw,kf. |
| 110 | (high* temperature* or extreme* temperature*).tw,kf. |
| 111 | (winter or summer or season*).tw,kf. |
| 112 | flood*.tw,kf. |
| 113 | (climate adj2 chang*).tw,kf. |
| 114 | (climate* adj2 event*).tw,kf. |
| 115 | (meteorological varia* or meteorological chang*).tw,kf. |
| 116 | rain*.tw,kf. |
| 117 | snow*.tw,kf. |
| 118 | drought*.tw,kf. |
| 119 | extreme cold.tw,kf. |
| 120 | freezing temperature*.tw,kf. |
| 121 | (storm or storms or rainstorm*).tw,kf. |
| 122 | sea-level*.tw,kf. |
| 123 | wildfire*.tw,kf. |
| 124 | (cyclon* or hurricane* or typhoon*).tw,kf. |
| 125 | (air pollut* or air quality).tw,kf. |
| 126 | pollen.tw,kf. |
| 127 | ozone.tw,kf. |
| 128 | particulate*.tw,kf. |
| 129 | emissions.tw,kf. |
| 130 | global warming.tw,kf. |
| 131 | greenhouse effect*.tw,kf. |
| 132 | heatwave*.tw,kf. |
| 133 | (extreme weather* or severe weather*).tw,kf. |
| 134 | extreme event*.tw,kf. |
| 135 | (weather-related or weather damag*).tw,kf. |
| 136 | Hot Temperature/ |
| 137 | Extreme Heat/ |
| 138 | Floods/ |
| 139 | Droughts/ |
| 140 | exp Climate Change/ |
| 141 | Greenhouse Effect/ |
| 142 | exp Rain/ |
| 143 | Meteorological Concepts/ |

| | |
|---|---|
| 144 | Extreme Weather/ |
| 145 | Extreme Cold Weather/ |
| 146 | Cyclonic Storms/ |
| 147 | Wildfires/ |
| 148 | Tornadoes/ |
| 149 | Tidal Waves/ |
| 150 | Landslides/ |
| 151 | exp Ozone/ |
| 152 | Particulate Matter/ |
| 153 | Air Pollution/ or Air Pollutants/ |
| 154 | Carbon Footprint/ |
| 155 | vector-borne disease*.tw,kf. |
| 156 | lyme disease.tw,kf. |
| 157 | yellow fever.tw,kf. |
| 158 | zika.tw,kf. |
| 159 | malaria.tw,kf. |
| 160 | (tick* or mosquito* or midge* or flea*).tw,kf. |
| 161 | airborne disease*.tw,kf. |
| 162 | aeroallergen*.tw,kf. |
| 163 | environmental degradation.tw,kf. |
| 164 | (greenspace* or green space*).tw,kf. |
| 165 | Vector-Borne Diseases/ |
| 166 | Tick-Borne Diseases/ |
| 167 | Lyme Disease/ |
| 168 | Malaria/ |
| 169 | Yellow Fever/ |
| 170 | Zika Virus Infection/ |
| 171 | Ticks/ |
| 172 | Culicidae/ or Siphonaptera/ |
| 173 | or/106-172 |
| 174 | exp Great Britain/ |
| 175 | (national health service* or nhs*).ti,ab,in. |
| 176 | (english not ((published or publication* or translat* or written or language* or speak* or literature or citation*) adj5 english)).ti,ab. |
| 177 | (gb or "g.b." or britain* or (british* not "british columbia") or uk or "u.k." or united kingdom* or (england* not "new england") or northern ireland* or northern irish* or scotland* or scottish* or ((wales or "south wales") not "new south wales") or welsh*).ti,ab,jw,in. |
| 178 | (bath or "bath's" or ((birmingham not alabama*) or ("birmingham's" not alabama*) or bradford or "bradford's" or brighton or "brighton's" or bristol or "bristol's" or carlisle* or "carlisle's" or (cambridge not (massachusetts* or boston* or harvard*)) or ("cambridge's" not (massachusetts* or boston* or harvard*)) or (canterbury not zealand*) or ("canterbury's" not zealand*) or chelmsford or "chelmsford's" or chester or "chester's" or chichester or "chichester's" or coventry or "coventry's" or derby or "derby's" or (durham not (carolina* or nc)) or ("durham's" not (carolina* or nc)) or ely or "ely's" or exeter or "exeter's" or gloucester or "gloucester's" or hereford or "hereford's" or hull or "hull's" or lancaster or "lancaster's" or leeds* or leicester or "leicester's" or (lincoln not nebraska*) or ("lincoln's" not nebraska*) or (liverpool not (new south wales* or nsw)) or ("liverpool's" not (new south wales* or nsw)) or ((london not (ontario* or ont or toronto*)) or ("london's" not (ontario* or ont or toronto*)) or manchester or "manchester's" or (newcastle not (new south wales* or nsw)) or ("newcastle's" not (new south wales* or nsw)) or norwich or "norwich's" or nottingham or |

| | |
|---|---|
| | "nottingham's" or oxford or "oxford's" or peterborough or "peterborough's" or plymouth or "plymouth's" or portsmouth or "portsmouth's" or preston or "preston's" or ripon or "ripon's" or salford or "salford's" or salisbury or "salisbury's" or sheffield or "sheffield's" or southampton or "southampton's" or st albans or stoke or "stoke's" or sunderland or "sunderland's" or truro or "truro's" or wakefield or "wakefield's" or wells or westminster or "westminster's" or winchester or "winchester's" or wolverhampton or "wolverhampton's" or (worcester not (massachusetts* or boston* or harvard*)) or ("worcester's" not (massachusetts* or boston* or harvard*)) or (york not ("new york*" or ny or ontario* or ont or toronto*)) or ("york's" not ("new york*" or ny or ontario* or ont or toronto*))))).ti,ab,in. |
| 179 | (bangor or "bangor's" or cardiff or "cardiff's" or newport or "newport's" or st asaph or "st asaph's" or st davids or swansea or "swansea's").ti,ab,in. |
| 180 | (aberdeen or "aberdeen's" or dundee or "dundee's" or edinburgh or "edinburgh's" or glasgow or "glasgow's" or inverness or (perth not australia*) or ("perth's" not australia*) or stirling or "stirling's").ti,ab,in. |
| 181 | (armagh or "armagh's" or belfast or "belfast's" or lisburn or "lisburn's" or londonderry or "londonderry's" or derry or "derry's" or newry or "newry's").ti,ab,in. |
| 182 | or/174-181 |
| 183 | (exp africa/ or exp americas/ or exp antarctic regions/ or exp arctic regions/ or exp asia/ or exp oceania/) not (exp great britain/ or europe/) |
| 184 | 182 not 183 |
| 185 | 105 and 173 |
| 186 | 184 and 185 |
| 187 | limit 186 to yr="2010 - 2023" |

**Ovid Embase**

| | |
|---|---|
| 1 | (deprivation or deprived or disadvantage* or vulnerable).tw,kf. |
| 2 | social risk*.tw,kf. |
| 3 | (sociodemographic* or socioeconomic*).tw,kf. |
| 4 | (inclusion health group* or socially excluded).tw,kf. |
| 5 | working class*.tw,kf. |
| 6 | poverty.tw,kf. |
| 7 | house* income*.tw,kf. |
| 8 | unemployed.tw,kf. |
| 9 | ethnic minorit*.tw,kf. |
| 10 | (Asian British or Black British or South Asian* or Afr* Caribbean*).tw,kf. |
| 11 | protected characteristic*.tw,kf. |
| 12 | (religion or belief*).tw,kf. |
| 13 | sexual orientation.tw,kf. |
| 14 | LGBTQ+.tw,kf. |
| 15 | (lesbian or gay).tw,kf. |
| 16 | (bisexual or intersex or queer or asexual or CIS or non-binary or pansexual).tw,kf. |
| 17 | gender diverse.tw,kf. |
| 18 | transgender.tw,kf. |
| 19 | (homeless* or rough sleep* or vagrant*).tw,kf. |
| 20 | (substance* adj2 (use* or abuse* or misuse or addict* or depend*)).tw,kf. |

| | |
|---|---|
| 21 | (drug* adj2 (use* or abuse* or misuse or addict* or depend*)).tw,kf. |
| 22 | (alcohol* adj2 (use* or abuse* or misuse or addict* or depend*)).tw,kf. |
| 23 | (alcoholic* or alcoholism).tw,kf. |
| 24 | (migrant* or immigrant*).tw,kf. |
| 25 | (gypsy or gypsies).tw,kf. |
| 26 | Roma.tw,kf. |
| 27 | (travelling communit* or traveller* or traveler*).tw,kf. |
| 28 | (sex work* or prostitut*).tw,kf. |
| 29 | justice system*.tw,kf. |
| 30 | modern slave*.tw,kf. |
| 31 | (forced labour or forced labor).tw,kf. |
| 32 | human trafficking.tw,kf. |
| 33 | enslavement.tw,kf. |
| 34 | (prisoner* or offend* or remand* or incarcarat* or imprisonment or custod*).tw,kf. |
| 35 | asylum seeker*.tw,kf. |
| 36 | refugee*.tw,kf. |
| 37 | displaced person*.tw,kf. |
| 38 | social deprivation/ |
| 39 | economic status/ or lowest income group/ |
| 40 | exp poverty/ |
| 41 | ethnic group/ |
| 42 | socioeconomics/ |
| 43 | Religion/ |
| 44 | minority group/ |
| 45 | cultural anthropology/ |
| 46 | exp "sexual and gender minority"/ |
| 47 | sexual characteristics/ |
| 48 | exp Homosexuality/ |
| 49 | homelessness/ |
| 50 | drug dependence/ |
| 51 | alcoholism/ |
| 52 | "drug use"/ |
| 53 | Human Trafficking/ |
| 54 | exp vulnerable population/ |
| 55 | migration/ |
| 56 | "romani (people)"/ |
| 57 | sex worker/ or prostitution/ |
| 58 | prisoner/ |
| 59 | crime victim/ |
| 60 | offender/ |
| 61 | exp migrant/ |
| 62 | slave/ |
| 63 | exp refugee/ |

| | |
|---|---|
| 64 | Working Poor/ |
| 65 | slavery/ |
| 66 | or/1-65 |
| 67 | (prison or prisons or jail* or gaol* or detention or correctional).tw,kf. |
| 68 | ((camp* or tent or tents) adj2 (transit* or temporary or pitch* or site*)).tw,kf. |
| 69 | campsite*.tw,kf. |
| 70 | ((asylum or migrant* or refugee* or immigrant* or immigration) adj2 (centre* or center* or camp* or support* or removal*)).tw,kf. |
| 71 | caravan*.tw,kf. |
| 72 | (park home* or mobile home*).tw,kf. |
| 73 | trailer park*.tw,kf. |
| 74 | outreach program*.tw,kf. |
| 75 | shelter*.tw,kf. |
| 76 | (temporary hous* or "no fixed abode").tw,kf. |
| 77 | temporary accommodation.tw,kf. |
| 78 | (supported hous* or social hous* or half-way house* or council hous*).tw,kf. |
| 79 | hostel*.tw,kf. |
| 80 | (food bank* or foodbank*).tw,kf. |
| 81 | soup kitchen*.tw,kf. |
| 82 | (rehabilitation centre* or rehabilitation centre*).tw,kf. |
| 83 | ((substance or drug* or alcohol*) and treatment clinic*).tw,kf. |
| 84 | (detoxification centre* or detoxification center*).tw,kf. |
| 85 | (drug* counsel* or alcohol* counsel*).tw,kf. |
| 86 | harm reduction program*.tw,kf. |
| 87 | sober living home*.tw,kf. |
| 88 | needle exchange program*.tw,kf. |
| 89 | methadone clinic*.tw,kf. |
| 90 | (GUM or genitourinary medicine clinic*).tw,kf. |
| 91 | (red light district or brothel*).tw,kf. |
| 92 | sexual health clinic*.tw,kf. |
| 93 | refugee camp/ |
| 94 | Emergency Shelter/ |
| 95 | exp correctional facility/ |
| 96 | detention center/ or asylum seeker center/ |
| 97 | housing/ |
| 98 | rehabilitation center/ |
| 99 | drug dependence treatment/ |
| 100 | Counseling/ |
| 101 | Harm Reduction/ |
| 102 | preventive health service/ |
| 103 | or/67-102 |
| 104 | 66 or 103 |
| 105 | climat*.tw,kf. |
| 106 | extreme heat.tw,kf. |

| | |
|---|---|
| 107 | (temperature adj increase*).tw,kf. |
| 108 | ambient temperature*.tw,kf. |
| 109 | (high* temperature* or extreme* temperature*).tw,kf. |
| 110 | (winter or summer or season*).tw,kf. |
| 111 | flood*.tw,kf. |
| 112 | (climate adj2 chang*).tw,kf. |
| 113 | (climate* adj2 event*).tw,kf. |
| 114 | (meteorological varia* or meteorological chang*).tw,kf. |
| 115 | rain*.tw,kf. |
| 116 | snow*.tw,kf. |
| 117 | drought*.tw,kf. |
| 118 | extreme cold.tw,kf. |
| 119 | freezing temperature*.tw,kf. |
| 120 | (storm or storms or rainstorm*).tw,kf. |
| 121 | sea-level*.tw,kf. |
| 122 | wildfire*.tw,kf. |
| 123 | (cyclon* or hurricane* or typhoon*).tw,kf. |
| 124 | (air pollut* or air quality).tw,kf. |
| 125 | pollen.tw,kf. |
| 126 | ozone.tw,kf. |
| 127 | particulate*.tw,kf. |
| 128 | emissions.tw,kf. |
| 129 | global warming.tw,kf. |
| 130 | greenhouse effect*.tw,kf. |
| 131 | heatwave*.tw,kf. |
| 132 | (extreme weather* or severe weather*).tw,kf. |
| 133 | extreme event*.tw,kf. |
| 134 | (weather-related or weather damag*).tw,kf. |
| 135 | high temperature/ |
| 136 | extreme hot weather/ |
| 137 | flooding/ |
| 138 | drought/ |
| 139 | exp Climate Change/ |
| 140 | Greenhouse Effect/ |
| 141 | exp Rain/ |
| 142 | meteorological phenomena/ |
| 143 | severe weather/ or extreme weather/ |
| 144 | Extreme Cold Weather/ |
| 145 | hurricane/ |
| 146 | exp wildfire/ |
| 147 | tornado/ |
| 148 | tsunami/ |
| 149 | landslide/ |

| | |
|---|---|
| 150 | ozone/ |
| 151 | exp particulate matter/ |
| 152 | air pollution/ or air pollutant/ |
| 153 | carbon footprint/ or greenhouse gas emission/ |
| 154 | vector-borne disease*.tw,kf. |
| 155 | lyme disease.tw,kf. |
| 156 | yellow fever.tw,kf. |
| 157 | zika.tw,kf. |
| 158 | malaria.tw,kf. |
| 159 | (tick* or mosquito* or midge* or flea*).tw,kf. |
| 160 | airborne disease*.tw,kf. |
| 161 | aeroallergen*.tw,kf. |
| 162 | environmental degradation.tw,kf. |
| 163 | (greenspace* or green space*).tw,kf. |
| 164 | vector borne disease/ |
| 165 | tick borne disease/ |
| 166 | Lyme Disease/ |
| 167 | malaria/ or mosquito borne disease/ |
| 168 | Yellow Fever/ |
| 169 | Zika fever/ |
| 170 | tick/ |
| 171 | mosquito/ or flea/ |
| 172 | or/105-171 |
| 173 | 104 and 172 |
| 174 | United Kingdom/ |
| 175 | (national health service* or nhs*).ti,ab,in,ad. |
| 176 | (english not ((published or publication* or translat* or written or language* or speak* or literature or citation*) adj5 english)).ti,ab. |
| 177 | (gb or "g.b." or britain* or (british* not "british columbia") or uk or "u.k." or united kingdom* or (england* not "new england") or northern ireland* or northern irish* or scotland* or scottish* or ((wales or "south wales") not "new south wales") or welsh*).ti,ab,jw,in,ad. |
| 178 | (bath or "bath's" or ((birmingham not alabama*) or ("birmingham's" not alabama*) or bradford or "bradford's" or brighton or "brighton's" or bristol or "bristol's" or carlisle* or "carlisle's" or (cambridge not (massachusetts* or boston* or harvard*)) or ("cambridge's" not (massachusetts* or boston* or harvard*)) or (canterbury not zealand*) or ("canterbury's" not zealand*) or chelmsford or "chelmsford's" or chester or "chester's" or chichester or "chichester's" or coventry or "coventry's" or derby or "derby's" or (durham not (carolina* or nc)) or ("durham's" not (carolina* or nc)) or ely or "ely's" or exeter or "exeter's" or gloucester or "gloucester's" or hereford or "hereford's" or hull or "hull's" or lancaster or "lancaster's" or leeds* or leicester or "leicester's" or (lincoln not nebraska*) or ("lincoln's" not nebraska*) or (liverpool not (new south wales* or nsw)) or ("liverpool's" not (new south wales* or nsw)) or ((london not (ontario* or ont or toronto*)) or ("london's" not (ontario* or ont or toronto*)) or manchester or "manchester's" or (newcastle not (new south wales* or nsw)) or ("newcastle's" not (new south wales* or nsw)) or norwich or "norwich's" or nottingham or "nottingham's" or oxford or "oxford's" or peterborough or "peterborough's" or plymouth or "plymouth's" or |

| | |
|---|---|
| | portsmouth or "portsmouth's" or preston or "preston's" or ripon or "ripon's" or salford or "salford's" or salisbury or "salisbury's" or sheffield or "sheffield's" or southampton or "southampton's" or st albans or stoke or "stoke's" or sunderland or "sunderland's" or truro or "truro's" or wakefield or "wakefield's" or wells or westminster or "westminster's" or winchester or "winchester's" or wolverhampton or "wolverhampton's" or (worcester not (massachusetts* or boston* or harvard*)) or ("worcester's" not (massachusetts* or boston* or harvard*)) or (york not ("new york*" or ny or ontario* or ont or toronto*)) or ("york's" not ("new york*" or ny or ontario* or ont or toronto*))))).ti,ab,in,ad. |
| 179 | (bangor or "bangor's" or cardiff or "cardiff's" or newport or "newport's" or st asaph or "st asaph's" or st davids or swansea or "swansea's").ti,ab,in,ad. |
| 180 | (aberdeen or "aberdeen's" or dundee or "dundee's" or edinburgh or "edinburgh's" or glasgow or "glasgow's" or inverness or (perth not australia*) or ("perth's" not australia*) or stirling or "stirling's").ti,ab,in,ad. |
| 181 | (armagh or "armagh's" or belfast or "belfast's" or lisburn or "lisburn's" or londonderry or "londonderry's" or derry or "derry's" or newry or "newry's").ti,ab,in,ad. |
| 182 | or/174-181 |
| 183 | (exp "arctic and antarctic"/ or exp oceanic regions/ or exp western hemisphere/ or exp africa/ or exp asia/) not (united kingdom/ or europe/) |
| 184 | 182 not 183 |
| 185 | 173 and 184 |
| 186 | limit 185 to yr="2010 - 2023" |
| 187 | limit 186 to conference abstracts |
| 188 | 186 not 187 |

**Web of Science**

| | |
|---|---|
| #1 | TS=(deprivation or deprived or disadvantage* or vulnerable) OR TI=("social risk*") OR TS=(sociodemographic* or socioeconomic*) OR TS=("inclusion health group*" or "socially excluded") OR TS=("working class*") OR TI=(poverty) OR TS=("house* income*") OR TI=(unemployed) OR TI=("ethnic minorit*") OR TS=("Asian British" or "Black British" or "South Asian*" or "Afr* Caribbean*") OR TS=("protected characteristic*") OR TI=(religion or belief*) OR TS=("sexual orientation") OR TS=(LGBTQ+) OR TS=(lesbian or gay) OR TS=(bisexual or intersex or queer or asexual or CIS or non-binary or pansexual) OR TS=("gender diverse") OR TS=(transgender) OR TI=(homeless* or "rough sleep*" or vagrant*) OR TI=(substance* NEAR/1 (use* or abuse* or misuse or addict* or depend*)) OR TI=(drug* NEAR/1 (use* or abuse* or misuse or addict* or depend*)) OR TI=(alcohol* NEAR/1 (use* or abuse* or misuse or addict* or depend*)) OR TI=(alcoholic* or alcoholism) OR TS=(migrant* or immigrant*) OR TS=(gypsy or gypsies) OR TS=(Roma) OR TS=("travelling communit*" or traveller* or traveler*) OR TS=("sex work*" or prostitut*) OR TS=("justice system*") OR TS=("modern slave*") OR TS=("forced labour" or "forced labor") OR TS=("human trafficking")OR TS=(enslavement) OR TS=(prisoner* or offend* or remand* or incarcarat* or imprisonment or custod*) OR TS=("asylum seeker*") OR TS=(refugee*) OR TS=("displaced person*") |

| | |
|---|---|
| #2 | TS=(prison or prisons or jail* or gaol* or detention or correctional) OR TS=((camp* or tent or tents) NEAR/1 (transit* or temporary or pitch* or site*)) OR TS=(campsite*) OR TS=((asylum or migrant* or refugee* or immigrant* or immigration) NEAR/1 (centre* or center* or camp* or support* or removal*)) OR TS=(caravan*) OR TS=("park home*" or "mobile home*") OR TS=("trailer park*") OR TS=("outreach program*") OR TS=(shelter*) OR TS=("temporary hous*" or "no fixed abode") OR TS=("temporary accommodation") OR TS=("supported hous*" or "social hous*" or "half-way house*" or "council hous*") OR TS=(hostel*) OR TS=("food bank*" or foodbank*) OR TS=("soup kitchen*") OR TS=("rehabilitation centre*" or "rehabilitation centre*") OR TS=((substance or drug* or alcohol*) and "treatment clinic*") OR TS=("detoxification centre*" or "detoxification center*") OR TS=("drug* counsel*" or "alcohol* counsel*") OR TS=("harm reduction program*") OR TS=("sober living home*") OR TS=("needle exchange program*") OR TS=("methadone clinic*") OR TS=(GUM or "genitourinary medicine clinic*") OR TS=("red light district" or brothel*) OR TS=("sexual health clinic*" |
| #3 | #2 OR #1 |
| #4 | TI=(climat*) OR TS=("climate chang*") OR TS=("extreme heat") OR TS=("temperature increase*") OR TS=("ambient temperature*") OR TS=("hot temperature*" or "extreme* temperature*") OR TI=(winter or summer or season*) OR TS=(flood*) OR TS=("climate* event*") OR TS=("meteorological varia*" or "meteorological chang*") OR TI=(rain*) OR TI=(snow*) OR TS=(drought*) OR TS=("extreme cold") OR TS=("freezing temperature*") OR TS=(storm or storms or rainstorm*) OR TS=("sea-level*") OR TS=(wildfire*) OR TS=(cyclon* or hurricane* or typhoon*) OR TS=("air pollut*" or "air quality") OR TS=(pollen) OR TS=(ozone) OR TS=(particulate*) OR TS=("greenhouse emission*") OR TS=("global warming") OR TS=("greenhouse effect*") OR TS=(heatwave*) OR TS=("extreme weather*" or "severe weather*") OR TS=("extreme event*") OR TS=("weather-related" or "weather damag*") OR TS=("vector-borne disease*") OR TS=("lyme disease") OR TS=("yellow fever") OR TS=(zika) OR TS=(malaria) OR TS=(tick* or mosquito* or midge* or flea*) OR TS=("airborne disease*") OR TS=(aeroallergen*) OR TS=("environmental degradation") OR TS=(greenspace* or "green space*") |
| #5 | #4 AND #3 |
| #6 | AD=("national health service*" OR NHS) OR TS=("national health service*" OR NHS) OR TS=(english NOT ((published or publication* or translat* or written or language* or speak* or literature or citation*) NEAR/5 english)) OR AD=(gb or "g.b." or britain* or (british* NOT "british columbia") or uk or "u.k." or united kingdom* or (england* NOT "new england") or northern ireland* or northern irish* or scotland* or scottish* or ((wales or "south wales") NOT "new south wales") or welsh*) OR TS=(gb or "g.b." or britain* or (british* not "british columbia") or uk or "u.k." or united kingdom* or (england* NOT "new england") or northern ireland* or northern irish* or scotland* or scottish* or ((wales or "south wales") NOT "new south wales") or welsh*) OR CU=(gb or "g.b." or britain* or (british* not "british columbia") or uk or "u.k." or united kingdom* or (england* NOT "new england") or northern ireland* or northern irish* or scotland* or scottish* or ((wales or "south wales") NOT "new south wales") or welsh*) OR AD=(bath or "bath's") OR TS=(bath or "bath's") OR CI=(bath or "bath's") OR AD=((birmingham NOT alabama*) or ("birmingham's" NOT alabama*)) OR TS=((birmingham NOT alabama*) or ("birmingham's" NOT alabama*)) OR CI=((birmingham NOT alabama*) or ("birmingham's" NOT alabama*)) OR AD=(bradford or "bradford's" or brighton or "brighton's" or bristol or "bristol's" or carlisle* or "carlisle's") OR TS=(bradford or "bradford's" or brighton or |

| | |
|---|---|
| | "brighton's" or bristol or "bristol's" or carlisle* or "carlisle's") OR CI=(bradford or "bradford's" or brighton or "brighton's" or bristol or "bristol's" or carlisle* or "carlisle's") OR AD=("cambridge's" NOT (massachusetts* or boston* or harvard*)) OR TS=("cambridge's" NOT (massachusetts* or boston* or Harvard)) OR CI=("cambridge's" NOT (massachusetts* or boston* or Harvard)) OR AD=((canterbury NOT zealand*) or ("canterbury's" NOT zealand*)) OR TS=((canterbury NOT zealand*) or ("canterbury's" NOT zealand*)) OR CI=((canterbury NOT zealand*) or ("canterbury's" NOT zealand*)) OR AD=(chelmsford or "chelmsford's" or chester or "chester's" or chichester or "chichester's" or coventry or "coventry's" or derby or "derby's”) OR TS=(chelmsford or "chelmsford's" or chester or "chester's" or chichester or "chichester's" or coventry or "coventry's" or derby or "derby's”) OR CI=(chelmsford or "chelmsford's" or chester or "chester's" or chichester or "chichester's" or coventry or "coventry's" or derby or "derby's”) OR AD=(durham NOT (carolina* or nc)) OR TS=(durham NOT (carolina* or nc)) OR CI=(durham NOT (carolina* or nc)) OR AD=("durham's" NOT (carolina* or nc)) OR TS=("durham's" NOT (carolina* or nc)) OR CI=("durham's" NOT (carolina* or nc)) OR AD=(ely or "ely's" or exeter or "exeter's" or gloucester or "gloucester's" or hereford or "hereford's" or hull or "hull's" or lancaster or "lancaster's" or leeds* or leicester or "leicester's") OR TS=(ely or "ely's" or exeter or "exeter's" or gloucester or "gloucester's" or hereford or "hereford's" or hull or "hull's" or lancaster or "lancaster's" or leeds* or leicester or "leicester's") OR CI=(ely or "ely's" or exeter or "exeter's" or gloucester or "gloucester's" or hereford or "hereford's" or hull or "hull's" or lancaster or "lancaster's" or leeds* or leicester or "leicester's") OR AD=((lincoln NOT nebraska*) or ("lincoln's" NOT nebraska*)) OR TS=((lincoln NOT nebraska*) or ("lincoln's" NOT nebraska*)) OR CI=((lincoln NOT nebraska*) or ("lincoln's" NOT nebraska*)) OR AD=((liverpool NOT (new south wales* or nsw)) or ("liverpool's" NOT (new south wales* or nsw))) OR TS=((liverpool NOT (new south wales* or nsw)) or ("liverpool's" NOT (new south wales* or nsw))) OR CI=((liverpool NOT (new south wales* or nsw)) or ("liverpool's" NOT (new south wales* or nsw))) OR AD=(london NOT (ontario* or ont or toronto*)) OR TS=(london NOT (ontario* or ont or toronto*)) OR CI=(london NOT (ontario* or ont or toronto*)) OR AD=("london's" NOT (ontario* or ont or toronto*)) OR TS=("london's" NOT (ontario* or ont or toronto*)) OR CI=("london's" NOT (ontario* or ont or toronto*)) OR AD=(manchester or "manchester's") OR TS=(manchester or "manchester's") OR CI=(manchester or "manchester's") OR AD=((newcastle NOT (new south wales* or nsw)) or ("newcastle's" NOT (new south wales* or nsw))) OR TS=((newcastle NOT (new south wales* or nsw)) or ("newcastle's" NOT (new south wales* or nsw))) OR CI=((newcastle NOT (new south wales* or nsw)) or ("newcastle's" NOT (new south wales* or nsw))) OR AD=(norwich or "norwich's" or nottingham or "nottingham's" or oxford or "oxford's" or peterborough or "peterborough's" or plymouth or "plymouth's" or portsmouth or "portsmouth's" or preston or "preston's" or ripon or "ripon's" or salford or "salford's" or salisbury or "salisbury's" or sheffield or "sheffield's" or southampton or "southampton's" or st albans or stoke or "stoke's" or sunderland or "sunderland's" or truro or "truro's" or wakefield or "wakefield's" or wells or westminster or "westminster's" or winchester or "winchester's" or wolverhampton or "wolverhampton's") OR TS=(norwich or "norwich's" or nottingham or "nottingham's" or oxford or "oxford's" or peterborough or "peterborough's" or plymouth or "plymouth's" or portsmouth or "portsmouth's" or preston or "preston's" or ripon or "ripon's" or salford or "salford's" or salisbury or "salisbury's" or sheffield or "sheffield's" or southampton or "southampton's" or st albans or stoke or "stoke's" or sunderland or "sunderland's" or truro or "truro's" or wakefield or "wakefield's" or wells or westminster or "westminster's" or winchester or "winchester's" or wolverhampton or "wolverhampton's") OR CI=(norwich or "norwich's" or nottingham or |

| | |
|---|---|
| | "nottingham's" or oxford or "oxford's" or peterborough or "peterborough's" or plymouth or "plymouth's" or portsmouth or "portsmouth's" or preston or "preston's" or ripon or "ripon's" or salford or "salford's" or salisbury or "salisbury's" or sheffield or "sheffield's" or southampton or "southampton's" or st albans or stoke or "stoke's" or sunderland or "sunderland's" or truro or "truro's" or wakefield or "wakefield's" or wells or westminster or "westminster's" or winchester or "winchester's" or wolverhampton or "wolverhampton's") OR AD=(worcester NOT (massachusetts* or boston* or harvard*)) OR TS=(worcester NOT (massachusetts* or boston* or harvard*)) OR CI=(worcester NOT (massachusetts* or boston* or harvard*)) OR AD=(("worcester's" NOT (massachusetts* or boston* or harvard*)) or (york NOT ("new york*" or ny or ontario* or ont or toronto*)) or ("york's" NOT ("new york*" or ny or ontario* or ont or toronto*))) OR TS=(("worcester's" NOT (massachusetts* or boston* or harvard*)) or (york NOT ("new york*" or ny or ontario* or ont or toronto*)) or ("york's" NOT ("new york*" or ny or ontario* or ont or toronto*))) OR CI=(("worcester's" NOT (massachusetts* or boston* or harvard*)) or (york NOT ("new york*" or ny or ontario* or ont or toronto*)) or ("york's" NOT ("new york*" or ny or ontario* or ont or toronto*))) OR AD=(bangor or "bangor's" or cardiff or "cardiff's" or newport or "newport's" or st asaph or "st asaph's" or st davids or swansea or "swansea's") OR TS=(bangor or "bangor's" or cardiff or "cardiff's" or newport or "newport's" or st asaph or "st asaph's" or st davids or swansea or "swansea's") OR CI=( bangor or "bangor's" or cardiff or "cardiff's" or newport or "newport's" or st asaph or "st asaph's" or st davids or swansea or "swansea's") OR AD=(aberdeen or "aberdeen's" or dundee or "dundee's" or edinburgh or "edinburgh's" or glasgow or "glasgow's" or inverness or (perth NOT australia*) or ("perth's" NOT australia*) or stirling or "stirling's") OR TS=(aberdeen or "aberdeen's" or dundee or "dundee's" or edinburgh or "edinburgh's" or glasgow or "glasgow's" or inverness or (perth NOT australia*) or ("perth's" NOT australia*) or stirling or "stirling's") OR CI=(aberdeen or "aberdeen's" or dundee or "dundee's" or edinburgh or "edinburgh's" or glasgow or "glasgow's" or inverness or (perth NOT australia*) or ("perth's" NOT australia*) or stirling or "stirling's") OR AD=(armagh or "armagh's" or belfast or "belfast's" or lisburn or "lisburn's" or londonderry or "londonderry's" or derry or "derry's" or newry or "newry's") OR TS=(armagh or "armagh's" or belfast or "belfast's" or lisburn or "lisburn's" or londonderry or "londonderry's" or derry or "derry's" or newry or "newry's") OR CI=(armagh or "armagh's" or belfast or "belfast's" or lisburn or "lisburn's" or londonderry or "londonderry's" or derry or "derry's" or newry or "newry's") |
| #7 | #5 AND #6 |
| #8 | #7 [limited to Index Date 2023-07-17 to 2024-02-19] |
| #9 | #7 and Article or Early Access or Review Article (Document Types) |

**FAIR database**

(climat* OR heat* OR temperature* OR flood*  OR rain* OR drought* OR cold* OR sea-level* OR wildfire* OR pollution OR “global warming" OR "greenhouse effect" OR "extreme weather" OR "carbon footprint") AND (uk OR england OR “united kingdom” OR britain OR british OR london OR wales OR scotland OR “northern ireland” or NHS)

**King’s Fund library**

(climat* OR heat* OR temperature* OR flood*  OR rain* OR drought* OR cold* OR sea-level* OR wildfire* OR pollution OR “global warming" OR "greenhouse effect" OR "extreme weather" OR "carbon footprint")

Supplementary Material 5. List of excluded studies

**Exclusion reason: wrong study design (n=33)**

| Author | Year | Title |
|---|---|---|
| Armstrong and others | 2014 | Conditional Poisson models: a flexible alternative to conditional logistic case cross-over analysis |
| Andrijevic and others | 2023 | Towards scenario representation of adaptive capacity for global climate change assessments |
| Ascione and others | 2022 | The trend of heat-related mortality in European cities |
| Bates and others | 2012 | The impact of climate change upon health and health inequalities in the North West of England |
| Bennett and Friel | 2014 | Impacts of climate change on inequities in child health |
| Brennan and others | 2020 | Preventative strategies and interventions to improve outcomes during heatwaves |
| Buchs and others | 2011 | Who bears the brunt? Distributional effects of climate change mitigation policies |
| Crandon and others | 2022 | The clinical implications of climate change for mental health |
| De Chavez and others | 2017 | Using environmental monitoring to complement in-depth qualitative interviews in cold homes research |
| Ferguson and others | 2021 | Systemic inequalities in indoor air pollution exposure in London, UK |
| Fitton and Moncaster | 2019 | Arguments for a co-production approach to community flood protection |
| Gasparrini and Armstrong | 2010 | Time series analysis on the health effects of temperature: Advancements and limitations |
| Hagg and others | 2019 | Scotland adapts: A capability framework for a climate ready public sector |
| Hosford and others | 2021 | The effects of road pricing on transportation and health equity: A scoping review |
| Jay and others | 2021 | Reducing the health effects of hot weather and heat extremes: from personal cooling strategies to green cities |
| Khosla and others | 2021 | Health risks of extreme heat |
| Kroeger | 2023 | Households worldwide become food insecure when it is too hot |
| Lindley and others | 2011 | Climate change, justice and vulnerability |
| McKee and others | 2021 | The changing health needs of the UK population |
| Munro and others | 2020 | Sustainable health equity: achieving a net-zero UK |
| Negev and Kovats | 2016 | Climate change adaptation in the reorganized UK public health system: a view from local government |
| NHS Providers | 2021 | Climate change is a public health emergency: the NHS is rising to the challenge |
| Oyebanjo and Bushell | 2014 | A critical evaluation of the UK SunSmart campaign and its relevance to Black and minority ethnic communities |

| Author | Year | Title |
| --- | --- | --- |
| Public Health England | 2018 | Heatwave plan for England: protecting health and reducing harm from extreme heat and heatwaves |
| Public Health Scotland | 2023 | Working together to build climate-resilient, healthy and equitable places A briefing for local government and partners |
| Ruane and others | 2016 | The Vulnerability, Impacts, Adaptation and Climate Services Advisory Board (VIACS AB v1.0) contribution to CMIP6 |
| Sayers and others | 2018 | Flood vulnerability, risk, and social disadvantage: current and future patterns in the UK |
| Slenning | 2010 | One health and climate change: linking environmental and animal health to human health |
| Stevens and others | 2022 | A comprehensive urban programme to reduce energy poverty and its effects on health and wellbeing of citizens in six European countries: study protocol of a controlled trial |
| Townend and others | 2021 | Operationalising coastal resilience to flood and erosion hazard: A demonstration for England |
| van Daalen and others | 2023 | Approaching unsafe limits: climate-related health inequities within and beyond Europe |
| Wan and others | 2023 | Heat-health governance in a cool nation: A case study of Scotland |
| Zaidi and Pelling | 2015 | Institutionally configured risk: Assessing urban resilience and disaster risk reduction to heat wave risk in London |

**Exclusion reason: wrong publication type (n=6)**

| Author | Year | Title |
| --- | --- | --- |
| Bone and others | 2010 | Will drivers for home energy efficiency harm occupant health? |
| Capone and others | 2023 | Interaction between air pollutants and pollen grains: effects on public and occupational health |
| Donaldson and Wedzicha | 2013 | Deprivation, winter season, and COPD exacerbations |
| Firth | 2014 | Healthy settings and developing wellbeing in the community: RSPH annual conference and awards ceremony – 1st October 2014 |
| Murray and Ebi | 2012 | IPCC special report on managing the risks of extreme events and disasters to advance climate change adaptation (SREX) |
| Rutter | 2019 | ‘July heatwave’ causes highest A&E demand ever |

**Exclusion reason: wrong language (n=1)**

| Author | Year | Title |
|---|---|---|
| Fernandez and Rodela | 2020 | “Hay poder en numeros”: understanding the development of a collectivist Latinx parent identity and conscientizacao amid an anti-immigrant climate |

**Exclusion reason: no explicit link to climate change (n=75)**

| Author | Year | Title |
|---|---|---|
| Al Ahad | 2022 | The spatial-temporal effect of air pollution on GP visits and hospital admissions by ethnicity in the United Kingdom: An individual-level analysis |
| Al Ahad | 2023 | The association of long-term exposure to outdoor air pollution with all-cause GP visits and hospital admissions by ethnicity and country of birth in the United Kingdom |
| Al Ahad and others | 2023 | The spatial-temporal effect of air pollution on individuals' reported health and its variation by ethnic groups in the United Kingdom: a multilevel longitudinal analysis |
| Al Ahad and others | 2022 | Does long-term air pollution exposure affect self-reported health and limiting long term illness disproportionately for ethnic minorities in the UK? A Census-based individual level analysis |
| Al Ahad and others | 2022 | Air pollution and individuals' mental wellbeing in the adult population in United Kingdom: A spatial-temporal longitudinal study and the moderating effect of ethnicity |
| Astell-Burt and others | 2013 | Effect of air pollution and racism on ethnic differences in respiratory health among adolescents living in an urban environment |
| Atkinson and others | 2013 | Long-term exposure to outdoor air pollution and incidence of cardiovascular diseases |
| Barnes and others | 2019 | Emissions vs exposure: Increasing injustice from road traffic-related air pollution in the United Kingdom |
| Bixby and others | 2015 | Associations between green space and health in English cities: an ecological, cross-sectional study |
| Brunt and others | 2017 | Air pollution, deprivation and health: understanding relationships to add value to local air quality management policy and practice in Wales, UK |
| Carey and others | 2018 | Are noise and air pollution related to the incidence of dementia? A cohort study in London, England |
| Charlton and others | 2023 | Long-term outdoor air pollution and COVID-19 mortality in London: an individual-level analysis |
| Chua and others | 2020 | Ambient air pollution associations with retinal morphology in the UK Biobank |

| Author | Year | Title |
| --- | --- | --- |
| Cruz and others | 2022 | Association of environmental and socioeconomic indicators with serious mental illness diagnoses identified from general practitioner practice data in England: A spatial Bayesian modelling study |
| Doiron and others | 2019 | Air pollution, lung function and COPD: results from the population-based UK Biobank study |
| Fecht and others | 2015 | Associations between air pollution and socioeconomic characteristics, ethnicity and age profile of neighbourhoods in England and the Netherlands |
| Feng and others | 2023 | The effects of long-term exposure to air pollution on incident mental disorders among patients with prediabetes and diabetes: Findings from a large prospective cohort |
| Ferguson and others | 2023 | Analysis of inequalities in personal exposure to PM2.5: A modelling study for the Greater London school-aged population |
| Gao and others | 2023 | Association between long-term exposure to wildfire-related PM2.5 and mortality: A longitudinal analysis of the UK Biobank |
| Geary and others | 2023 | Ambient greenness, access to local green spaces, and subsequent mental health: a 10-year longitudinal dynamic panel study of 2.3 million adults in Wales |
| Geary and others | 2023 | Green-blue space exposure changes and impact on individual-level well-being and mental health: a population-wide dynamic longitudinal panel study with linked survey data |
| Gray and others | 2023 | Deprivation based inequality in NOx emissions in England |
| Grey and others | 2017 | The short-term health and psychosocial impacts of domestic energy efficiency investments in low-income areas: a controlled before and after study |
| Halonen and others | 2016 | Long-term exposure to traffic pollution and hospital admissions in London |
| Hao and others | 2023 | Ethnic disparities in ambient air and traffic-related pollution exposure and ethnic-specific impacts on clinical biomarker levels |
| Hart and others | 2013 | Ambient air pollution exposures and risk of rheumatoid arthritis |
| Heyman and others | 2011 | A randomised controlled trial of an energy efficiency intervention for families living in fuel poverty |
| Horton and others | 2023 | Air pollution and public health vulnerabilities, susceptibilities and inequalities in Wales, UK |
| Jephcote and Chen | 2012 | Environmental injustices of children's exposure to air pollution from road-transport within the model British multicultural city of Leicester: 2000-09 |
| Johnes and others | 2023 | Using sensor data to identify factors affecting internal air quality within 279 lower income households in Cornwall, South West of England |

| Author | Year | Title |
|---|---|---|
| Karamanos and others | 2021 | Air pollution and trajectories of adolescent conduct problems: the roles of ethnicity and racism; evidence from the DASH longitudinal study |
| Karamanos and others | 2023 | Associations between air pollutants and blood pressure in an ethnically diverse cohort of adolescents in London, England |
| Kazakos and others | 2020 | Quantifying the health burden misclassification from the use of different PM2.5 exposure tier models: A case study of London |
| Kazakos and others | 2021 | Impact of COVID-19 lockdown on NO2 and PM2.5 exposure inequalities in London, UK |
| Keidel and others | 2019 | The role of socioeconomic status in the association of lung function and air pollution-A pooled analysis of 3 adult ESCAPE cohorts |
| Kelly and others | 2011 | The London low emission zone baseline study |
| Khan and others | 2018 | Criegee intermediates and their impacts on the troposphere |
| Kotecha and others | 2020 | Differential association of air pollution exposure with neonatal and postneonatal mortality in England and Wales: A cohort study |
| Laborde and others | 2011 | Assessment of training needs for disaster mental health preparedness in black communities |
| Lavigne and others | 2019 | Associations between metal constituents of ambient particulate matter and mortality in England: an ecological study |
| Lee and others | 2022 | Quantifying the impact of air pollution on COVID-19 hospitalisation and death rates in Scotland |
| Li and others | 2023 | Long-term exposure to air pollution and incident non-alcoholic fatty liver disease and cirrhosis: A cohort study |
| Li and others | 2023 | Associations of long-term joint exposure to various ambient air pollutants with all-cause and cause-specific mortality: evidence from a large population-based cohort study |
| Liu and others | 2024 | Exposure to residential green and blue space and the natural environment is associated with a lower incidence of psychiatric disorders in middle-aged and older adults: findings from the UK Biobank |
| Luo and others | 2022 | Long-term exposure to ambient air pollution is a risk factor for trajectory of cardiometabolic multimorbidity: A prospective study in the UK Biobank |
| Ma and others | 2023 | Exposure to various ambient air pollutants and 9 cardiovascular conditions among individuals with diabetes: A prospective analysis of the UK Biobank |
| Marks and others | 2016 | Geographical and temporal trends in imported infections from the tropics requiring inpatient care at the Hospital for Tropical Diseases, London - a 15 year study |
| Martin-Bassols and others | 2023 | Effect of in utero exposure to air pollution on adulthood hospitalizations |
| Mebrahtu and others | 2023 | Differences in public's perception of air quality and acceptability of a clean air zone: A mixed-methods cross sectional study |

| Author | Year | Title |
|---|---|---|
| Milojevic and others | 2017 | Socioeconomic and urban-rural differentials in exposure to air pollution and mortality burden in England |
| Mitchell and others | 2011 | A comparison of green space indicators for epidemiological research |
| Moran and others | 2022 | Does prison location matter for prisoner wellbeing? The effect of surrounding greenspace on self-harm and violence in prisons in England and Wales |
| Moran and others | 2023 | Greenspace in prison improves well-being irrespective of prisoner characteristics, with particularly beneficial effects for younger and unsentenced prisoners, and in overcrowded prisons |
| Morrison and others | 2014 | An initial assessment of spatial relationships between respiratory cases, soil metal content, air quality and deprivation indicators in Glasgow, Scotland, UK: relevance to the environmental justice agenda |
| Moyo and others | 2019 | Persistence of imported malaria Into the United Kingdom: An epidemiological review of risk factors and at-risk groups |
| Nachman and Parker | 2012 | Exposures to fine particulate air pollution and respiratory outcomes in adults using 2 national datasets: a cross-sectional study |
| Oliver and others | 2024 | A cross-sectional analysis of biodiversity, publicly accessible green space and mental well-being in Wales using routinely collected data |
| Raaschou-Nielsen and others | 2013 | Air pollution and lung cancer incidence in 17 European cohorts: Prospective analyses from the European Study of Cohorts for Air Pollution Effects (ESCAPE) |
| Richardson and others | 2013 | A regional measure of neighborhood multiple environmental deprivation: Relationships with health and health inequalities |
| Riddell and Babiker | 2017 | Imported dengue fever in East London: a 6-year retrospective observational study |
| Roca-Barcelo and others | 2020 | Risk of respiratory hospital admission associated with modelled concentrations of Aspergillus fumigatus from composting facilities in England |
| Ronaldson and others | 2022 | Associations between air pollution and multimorbidity in the UK Biobank: A cross-sectional study |
| Schembari and others | 2015 | Ambient air pollution and newborn size and adiposity at birth: Differences by maternal ethnicity (the Born in Bradford Study Cohort) |
| Sharpe and others | 2015 | Fuel poverty increases risk of mould contamination, regardless of adult risk perception & ventilation in social housing properties |
| Shi and others | 2023 | Dynamic association of ambient air pollution with incidence and mortality of pulmonary hypertension: A multistate trajectory analysis |
| Smith and others | 2019 | Characteristics of the environment and physical activity in midlife: Findings from UK Biobank |
| Temam and others | 2017 | Socioeconomic position and outdoor nitrogen dioxide (NO2) exposure in Western Europe: A multi-city analysis |

| Author | Year | Title |
|---|---|---|
| Tieges and others | 2022 | Investigating the association between regeneration of urban blue spaces and risk of incident chronic health conditions stratified by neighbourhood deprivation: A population-based retrospective study, 2000-2018 |
| Tonne and others | 2018 | Socioeconomic and ethnic inequalities in exposure to air and noise pollution in London |
| Tonne and Wilkinson | 2013 | Long-term exposure to air pollution is associated with survival following acute coronary syndrome |
| Travaglio and others | 2021 | Links between air pollution and COVID-19 in England |
| Wan and others | 2022 | Greenspace and mortality in the U.K. Biobank: Longitudinal cohort analysis of socio-economic, environmental, and biomarker pathways |
| Ward Thompson and others | 2016 | Mitigating stress and supporting health in deprived urban communities: The importance of green space and the social environment |
| Xie and others | 2021 | Interactions with artificial water features: A scoping review of health-related outcomes |
| Yap and others | 2012 | Association between long-term exposure to air pollution and specific causes of mortality in Scotland |

**Exclusion reason: wrong country (n=40)**

| Author | Year | Title |
|---|---|---|
| Adeleye and Tiwari | 2024 | Empirical assessment of methane emissions, socioeconomic factors, and infant mortality in Europe |
| Akritidis and others | 2024 | Strong increase in mortality attributable to ozone pollution under a climate change and demographic scenario |
| Bachmeyer and others | 2020 | Cases of malaria in travellers with sickle cell disease - Chemoprophylaxis is important for this risk group |
| Bai and others | 2023 | Neighborhood deprivation and rurality associated with patient-reported outcomes and survival in men with prostate cancer in NRG Oncology RTOG 0415 |
| Cassidy and others | 2024 | Regional temperature extremes and vulnerability under net zero $CO_2$ emissions |
| Chen and others | 2023 | Long-term exposure to low-level PM2.5 and mortality: Investigation of heterogeneity by harmonizing analyses in large cohort studies in Canada, United States, and Europe |
| Chen and others | 2024 | The impact of geopolitical risk on CO2 emissions inequality: Evidence from 38 developed and developing economies |
| Colón-González and others | 2021 | Projecting the risk of mosquito-borne diseases in a warmer and more populated world: a multi-model, multi-scenario intercomparison modelling study |

| Author | Year | Title |
|---|---|---|
| Conlon and others | 2020 | Mapping human vulnerability to extreme heat: A critical assessment of heat vulnerability indices created using principal components analysis |
| Doran and others | 2016 | Homelessness and other social determinants of health among emergency department patients |
| Ferrreira and others | 2019 | Home-based and informal work exposes the families to high levels of potentially toxic elements |
| Fewster and others | 2022 | Imminent loss of climate space for permafrost peatlands in Europe and Western Siberia |
| Gasparrini and others | 2017 | Projections of temperature-related excess mortality under climate change scenarios |
| Gebhardt and others | 2023 | The relationship of climate change awareness and psychopathology in persons with pre-existing mental health diagnoses |
| Harrington and Otto | 2023 | Underestimated climate risks from population ageing |
| Heltberg and Bonch-Osmolovskiy | 2011 | Mapping vulnerability to climate change - Mapping vulnerability to climate change |
| Khan and others | 2021 | Towards an efficient storm surge and inundation forecasting system over the Bengal delta: chasing the Supercyclone Amphan |
| Kihal-Talantikite and others | 2017 | Developing a data-driven spatial approach to assessment of neighbourhood influences on the spatial distribution of myocardial infarction |
| Kimutai and others | 2022 | Attribution of the human influence on heavy rainfall associated with flooding events during the 2012, 2016, and 2018 March-April-May seasons in Kenya |
| Kroeger | 2023 | Heat is associated with short-term increases in household food insecurity in 150 countries and this is mediated by income |
| Laryea and others | 2023 | Climate justice implications of banning air-freighted fresh produce |
| Leng and others | 2023 | Global inequities in population exposure to urban greenspaces increased amidst tree and nontree vegetation cover expansion |
| Li and others | 2023 | Green physical activity for leisure connects perceived residential greenspace and mental well-being |
| Little and others | 2023 | Future increased risk from extratropical windstorms in northern Europe |
| Lloyd and others | 2023 | The direct and indirect influences of interrelated regional-level sociodemographic factors on heat-attributable mortality in Europe: Insights for adaptation strategies |
| Lyons and others | 2023 | The effect of "smoky" coal bans on chronic lung disease among older people in Ireland |
| McDuffie and others | 2023 | The social cost of ozone-related mortality impacts from methane emissions |
| Moradi and others | 2023 | Particulate matter pollution remains a threat for cardiovascular health: Findings from the Global Burden of Disease 2019 |

| Author | Year | Title |
| --- | --- | --- |
| Moulds and others | 2021 | Modeling the impacts of urban flood risk management on social inequality |
| Reddington and others | 2023 | Inequalities in air pollution exposure and attributable mortality in a low carbon future |
| Ribeiro and others | 2015 | Development of a measure of multiple physical environmental deprivation. After United Kingdom and New Zealand, Portugal |
| Richardson and others | 2013 | Particulate air pollution and health inequalities: a Europe-wide ecological analysis |
| Rohat and others | 2019 | Influence of changes in socioeconomic and climatic conditions on future heat-related health challenges in Europe |
| Sunikka-Blank and Galvin | 2021 | Single parents in cold homes in Europe: How intersecting personal and national characteristics drive up the numbers of these vulnerable households |
| Tillett | 2011 | Pregnancy pause: Extreme heat linked to shortened gestation |
| Watts and others | 2015 | Health and climate change: Policy responses to protect public health |
| Wu and others | 2023 | From quantity to quality: Effects of urban greenness on life satisfaction and social inequality |
| Zhao and others | 2019 | Assessing socio-economic drought evolution characteristics and their possible meteorological driving force |
| Zhao and others | 2024 | Long-term prediction of the effects of climate change on indoor climate and air quality |
| Zhou and others | 2023 | The effects of heatwave on cognitive impairment among older adults: Exploring the combined effects of air pollution and green space |

**Exclusion reason: wrong population (n=137)**

| Author | Year | Title |
| --- | --- | --- |
| Ahmed and others | 2021 | Forecasting underheating in dwellings to detect excess winter mortality risks using time series models |
| Akram and Arabi | 2023 | Water-energy nexus for Birmingham, UK |
| Almendra and others | 2019 | Cold-related mortality in 3 European metropolitan areas: Athens, Lisbon and London. Implications for health promotion |
| Alves and others | 2018 | Multi-criteria approach for selection of green and grey infrastructure to reduce flood risk and increase co-benefits |
| Anonymous | 2016 | Heatwave plan outlines how staff can flag up risks to public health |
| Arbuthnott and others | 2018 | What is cold-related mortality? A multi-disciplinary perspective to inform climate change impact assessments |
| Arbuthnott and others | 2020 | Years of life lost and mortality due to heat and cold in the 3 largest English cities |

| Author | Year | Title |
|---|---|---|
| Armstrong and others | 2010 | Association of mortality with high temperatures in a temperate climate: England and Wales |
| Armstrong and others | 2018 | The impact of home energy efficiency interventions and winter fuel payments on winter- and cold-related mortality and morbidity in England: a natural equipment mixed-methods study |
| Ayling and others | 2021 | Impact of reduced rainfall on above ground dry matter production of semi-natural grassland in South Gloucestershire, UK: A rainfall manipulation study |
| Bei and others | 2013 | A prospective study of the impact of floods on the mental and physical health of older adults |
| Bhaskaran and others | 2010 | Short term effects of temperature on risk of myocardial infarction in England and Wales: time series regression analysis of the Myocardial Ischaemia National Audit Project (MINAP) registry |
| Bhaskaran and others | 2012 | Heat and risk of myocardial infarction: hourly level case-crossover analysis of MINAP database |
| Bohm and others | 2023 | Emotional reactions to climate change: a comparison across France, Germany, Norway, and the United Kingdom |
| Bolt and others | 2023 | Seasonality of acute kidney injury phenotypes in England: an unsupervised machine learning classification study of electronic health records |
| Bouzid and others | 2014 | Climate change and the emergence of vector-borne diseases in Europe: case study of dengue fever |
| Broderick and others | 2015 | Clinical, geographical, and temporal risk factors associated with presentation and outcome of vivax malaria imported into the United Kingdom over 27 years: observational study |
| Bryan and others | 2020 | The health and well-being effects of drought: assessing multi-stakeholder perspectives through narratives from the UK |
| Button and Coote | 2016 | Public health in a changing climate |
| Carroll and others | 2010 | Health and social impacts of a flood disaster: responding to needs and implications for practice |
| Chandwani and others | 2023 | Impact of environmental exposures on lung cancer in patients who never smoked |
| Chen and others | 2023 | The association between exposure to air pollution and dementia incidence: The modifying effect of smoking |
| Chen and others | 2023 | Ambient air pollution and risk of enterotomy, gastrointestinal cancer, and all-cause mortality among 4,708 individuals with inflammatory bowel disease: A prospective cohort study |
| Christidis and others | 2010 | Causes for the recent changes in cold- and heat-related mortality in England and Wales |

| Author | Year | Title |
|---|---|---|
| Christie and others | 2016 | Private needs, public responses: vulnerable people's flood-disrupted mobility |
| Clarke and others | 2021 | Inventories of extreme weather events and impacts: Implications for loss and damage from and adaptation to climate extremes |
| Climate Just | | Case study: Retrofitting UK hospitals to reduce overheating risk Academic case study of Addenbrooke Hospital, Cambridge |
| Coles and others | 2017 | Beyond 'flood hotspots': Modelling emergency service accessibility during flooding in York, UK |
| Committee on Climate Change | 2017 | UK climate change risk assessment 2017 synthesis report: priorities for the next five years |
| Cowen | 2012 | Hot weather increases death risk in psychosis patients |
| Dadvand and others | 2011 | Association between maternal exposure to ambient air pollution and congenital heart disease: A register-based spatiotemporal analysis |
| De'Donato and others | 2015 | Changes in the effect of heat on mortality in the last 20 years in nine European cities. Results from the PHASE Project |
| Emerson and others | 2019 | Risk of exposure to air pollution among British children with and without intellectual disabilities |
| Erens and others | 2021 | Public attitudes to, and behaviours taken during, hot weather by vulnerable groups: results from a national survey in England |
| Fernando and others | 2013 | Non-cirrhotic portal hypertension in the HIV-infected individual |
| Findlater and others | 2023 | Help-seeking following a flooding event: a cross-sectional analysis of adults affected by flooding in England in winter 2013/14 |
| Fontalba-Navas and others | 2017 | Incidence and risk factors for post-traumatic stress disorder in a population affected by a severe flood |
| Foster and others | 2021 | An advanced empirical model for quantifying the impact of heat and climate change on human physical work capacity |
| Fox-Rogers and others | 2016 | Is there really "nothing you can do"? Pathways to enhanced flood-risk preparedness |
| Gale and others | 2020 | Association between exposure to air pollution and prefrontal cortical volume in adults: A cross-sectional study from the UK biobank |
| Gasparrini and others | 2012 | The effect of high temperatures on cause-specific mortality in England and Wales |
| Glasper | 2011 | Planning for a heat wave: the implications for health care |
| Goldney and others | 2023 | Long-term ambient air pollution exposure and prospective change in sedentary behaviour and physical activity in individuals at risk of type 2 diabetes in the UK |
| Gough and others | 2019 | Assessment of overheating risk in gynaecology scanning rooms during near-heatwave conditions: A case study of the Royal Berkshire Hospital in the UK |

| Author | Year | Title |
|---|---|---|
| Graham and others | 2019 | Flood- and weather-damaged homes and mental health: An analysis using England's Mental Health Survey |
| Green and others | 2016 | Mortality during the 2013 heatwave in England--How did it compare to previous heatwaves? A retrospective observational study |
| Green and others | 2017 | City-scale accessibility of emergency responders operating during flood events |
| Grobusch and others | 2021 | Travel-related infections presenting in Europe: A 20-year analysis of EuroTravNet surveillance data |
| Hajat and Gasparrini | 2016 | The excess winter deaths measure why its use is misleading for public health understanding of cold-related health impacts |
| Hajat and others | 2016 | Public health vulnerability to wintertime weather: time-series regression and episode analyses of national mortality and morbidity databases to inform the Cold Weather Plan for England |
| Hajat and others | 2014 | Climate change effects on human health: projections of temperature-related mortality for the UK during the 2020s, 2050s and 2080s |
| Halliday and others | 2022 | The island effect: Spatial effects on mental wellbeing and residence on remote Scottish islands |
| Hamilton and others | 2015 | Health effects of home energy efficiency interventions in England: a modelling study |
| Hammer and others | 2023 | Assessment of the association between ambient air pollution and stillbirth in the UK: Results from a secondary analysis of the MiNESS case-control study |
| Hannam and others | 2014 | Air pollution exposure and adverse pregnancy outcomes in a large UK birth cohort: use of a novel spatio-temporal modelling technique |
| Health Protection Agency | 2012 | Health effects of climate change in the UK 2012 |
| Huang and others | 2020 | Weather regimes and patterns associated with temperature-related excess mortality in the UK: a pathway to sub-seasonal risk forecasting |
| Hughes and others | 2014 | Using an emergency department syndromic surveillance system to investigate the impact of extreme cold weather events |
| Jenkins and others | 2014 | Probabilistic spatial risk assessment of heat impacts and adaptations for London |
| Jenkins and others | 2022 | Updated projections of UK heat-related mortality using policy-relevant global warming levels and socio-economic scenarios |
| Jermacane and others | 2018 | The English national cohort study of flooding and health: the change in the prevalence of psychological morbidity at year 2 |
| Jiang and others | 2023 | Co-exposure to multiple air pollutants, genetic susceptibility, and the risk of myocardial infarction onset: A cohort analysis of the UK Biobank participants |

| Author | Year | Title |
|---|---|---|
| Johnson and Yu | 2020 | From flooding to finance: NHS ambulance-assisted evacuations of care home residents in Norfolk and Suffolk, UK |
| Kantamaneni and others | 2019 | Assessing and mapping regional coastal vulnerability for port environments and coastal cities |
| Kaye and others | 2023 | The impact of climate change and natural climate variability on the global distribution of Aedes aegypti |
| Killeen and others | 2017 | Measuring, manipulating and exploiting behaviours of adult mosquitoes to optimise malaria vector control impact |
| Killip and others | 2014 | Innovation in low-energy residential renovation: UK and France |
| Kim and Lee | 2019 | Differential mechanisms of potato yield loss induced by high day and night temperatures during tuber initiation and bulking: Photosynthesis and tuber growth |
| King | 2013 | Neighborhood walkable urban form and C-reactive protein |
| Konstantinoudis and others | 2023 | Asthma hospitalisations and heat exposure in England: a case-crossover study during 2002-2019 |
| Koscikova and Krivstov | 2023 | Environmental and social benefits of extensive green roofs applied on bus shelters in Edinburgh |
| Lelieveld and others | 2023 | Air pollution deaths attributable to fossil fuels: observational and modelling study |
| Li and others | 2019 | Lyme disease risks in Europe under multiple uncertain drivers of change |
| Li and others | 2023 | Long-term exposure to ambient air pollution, genetic susceptibility, and the incidence of bipolar disorder: A prospective cohort study |
| Liu and others | | Association between birth weight/joint exposure to ambient air pollutants and type 2 diabetes: a cohort study in the UK Biobank |
| Lomas and Kane | 2013 | Summertime temperatures and thermal comfort in UK homes |
| Lowe and others | 2016 | Evaluation of an early-warning system for heat wave-related mortality in Europe: Implications for sub-seasonal to seasonal forecasting and climate services |
| Luo and others | 2023 | Air pollution and allergic rhinitis: Findings from a prospective cohort study |
| Ma and others | 2024 | Genetic susceptibility modifies relationships between air pollutants and stroke risk: A large cohort study |
| Macintyre and others | 2023 | Impacts of emissions policies on future UK mortality burdens associated with air pollution |
| Madaniyazi and others | 2024 | Seasonality of mortality under climate change: a multicountry projection study |
| Mahmood and others | 2017 | Impact of air temperature on London Ambulance call-out incidents and response times |

| Author | Year | Title |
|---|---|---|
| Marques and others | 2017 | Cyclosporiasis in travellers returning to the United Kingdom from Mexico in summer 2017: lessons from the recent past to inform the future |
| Mason and others | 2010 | The psychological impact of exposure to floods |
| Masselot and others | 2023 | Excess mortality attributed to heat and cold: a health impact assessment study in 854 cities in Europe |
| Mavrogianni and others | 2010 | London housing and climate change: Impact on comfort and health – preliminary results of a summer overheating study |
| Mavrogianni and others | 2015 | Urban social housing resilience to excess summer heat |
| McRobert | 2010 | Flooding and the role of the local authority |
| Moss and others | 2022 | Spatio-temporal epidemiology of SARS-CoV-2 virus lineages in Teesside, UK, in 2020: effects of socio-economic deprivation, weather, and lockdown on lineage dynamics |
| Mulchandani and others | 2019 | Effect of insurance-related factors on the association between flooding and mental health outcomes |
| Mulchandani and others | 2020 | The English national cohort study of flooding & health: psychological morbidity at 3 years of follow up |
| Murage and others | 2017 | Effect of night-time temperatures on cause and age-specific mortality in London |
| Murage and others | 2018 | Variation in cold-related mortality in England since the introduction of the Cold Weather Plan: Which areas have the greatest unmet needs? |
| Nichols and Richardson | 2011 | Climate change, health and sustainability: a brief survey of primary care trusts in the south west of England |
| Ntontis and others | 2020 | Endurance or decline of emergent groups following a flood disaster: Implications for community resilience |
| Ntontis and others | 2021 | Collective resilience in the disaster recovery period: Emergent social identity and observed social support are associated with collective efficacy, well-being, and the provision of social support |
| Ogunbode and others | 2019 | The resilience paradox: flooding experience, coping and climate change mitigation intentions |
| Paranjothy and others | 2011 | Psychosocial impact of the summer 2007 floods in England |
| Parsons and others | 2010 | Modelling the effects of the weather on admissions to UK trauma units: a cross-sectional study |
| Pinsent and others | 2014 | Risk factors for UK Plasmodium falciparum cases |
| Prichard and others | 2022 | Differential health responses to climate change projections in 3 UK cities as measured by ambulance dispatch data |
| Psistaki and others | 2020 | Weather patterns and all-cause mortality in England, UK |
| Psistaki and Paschalidou | 2023 | The effect of apparent temperature on all-cause mortality in England, UK |

| Author | Year | Title |
|---|---|---|
| Rendell and others | 2020 | Public health implications of solar UV exposure during extreme cold and hot weather episodes in 2018 in Chilton, South East England |
| Robin and others | 2020 | Impact of flooding on health-related quality of life in England: results from the National Study of Flooding and Health |
| Rodopoulou and others | 2015 | Searching for the best modeling specification for assessing the effects of temperature and humidity on health: a time series analysis in 3 European cities |
| Rustemeyer and Howells | 2021 | Excess Mortality in England during the 2019 Summer Heatwaves |
| Sahani and others | 2022 | Heat risk of mortality in 2 different regions of the United Kingdom |
| Sartini and others | 2016 | Effect of cold spells and their modifiers on cardiovascular disease events: Evidence from 2 prospective studies |
| Scarborough and others | 2012 | Contribution of climate and air pollution to variation in coronary heart disease mortality rates in England |
| Seklecka and others | 2017 | Mortality effects of temperature changes in the United Kingdom |
| Seklecka and others | 2017 | Mortality effects of temperature changes in the United Kingdom |
| Sharpe and others | 2015 | Higher energy efficient homes are associated with increased risk of doctor diagnosed asthma in a UK subpopulation |
| Sharpe and others | 2019 | Household energy efficiency and health: Area-level analysis of hospital admissions in England |
| Sheffield and others | 2018 | Association between particulate air pollution exposure during pregnancy and postpartum maternal psychological functioning |
| Singh and others | 2024 | Impacts of ambient air quality on acute asthma hospital admissions during the COVID-19 pandemic in Oxford City, UK: a time-series study |
| Symonds and others | 2019 | MicroEnv: A microsimulation model for quantifying the impacts of environmental policies on population health and health inequalities |
| Syukrowardi and others | 2014 | Factors affecting resilience in elementary school-aged children after exposing the floods |
| Taylor and others | 2015 | Mapping the effects of urban heat island, housing, and age on excess heat-related mortality in London |
| Taylor and others | 2016 | Mapping indoor overheating and air pollution risk modification across Great Britain: A modelling study |
| Taylor and others | 2018 | Estimating the influence of housing energy efficiency and overheating adaptations on heat-related mortality in the West Midlands, UK |
| Taylor and others | 2021 | Projecting the impacts of housing on temperature-related mortality in London during typical future years |
| Tempest and others | 2017 | Secondary stressors are associated with probable psychological morbidity after flooding: a cross-sectional analysis |
| Tobías and others | 2012 | A cautionary note to prevent the heat effects on human health |

| Author | Year | Title |
|---|---|---|
| Tomlinson and others | 2011 | Including the urban heat island in spatial heat health risk assessment strategies: a case study for Birmingham, UK |
| Twiddy and others | 2022 | Understanding the long-term impact of flooding on the wellbeing of residents: A mixed methods study |
| Undorf and others | 2020 | Learning from the 2018 heatwave in the context of climate change: are high-temperature extremes important for adaptation in Scotland? |
| Vinten and others | 2019 | Water for all: Towards an integrated approach to wetland conservation and flood risk reduction in a lowland catchment in Scotland |
| Waite and others | 2017 | The English national cohort study of flooding and health: cross-sectional analysis of mental health outcomes at year one |
| Whittle and others | 2010 | Flood, vulnerability and urban resilience: a real-time study of local recovery following the floods of June 2007 in Hull |
| Whittle and others | 2010 | After the rain - learning the lessons from flood recovery in Hull. Final project report for 'Flood, Vulnerability and Urban Resilience: a real-time study of local recovery following the floods of June 2007 in Hull' |
| Wind and Komproe | 2012 | The mechanisms that associate community social capital with post-disaster mental health: A multilevel model |
| Wingfield and Brisley | 2017 | Assessment of the impact of recently built flood alleviation schemes in managing long-term residual flood risk in England |
| Wolf and others | 2014 | Performance assessment of a heat wave vulnerability index for Greater London, United Kingdom |
| Wong and others | 2018 | Physical, psychological, and social health impact of temperature rise due to urban heat island phenomenon and its associated factors |
| Yi and others | 2023 | Modelling urban dwellers' indoor heat stress to enhance heat-health warning and planning |
| Zhang and others | 2023 | Assessment of short-term heat effects on cardiovascular mortality and vulnerability factors using small area data in Europe |

**Exclusion reason: wrong exposure (n=24)**

| Author | Year | Title |
|---|---|---|
| Astell-Burt and others | 2014 | The association between green space and mental health varies across the lifecourse. A longitudinal study |
| Bakolis and others | 2021 | Mental health consequences of urban air pollution: prospective population-based longitudinal survey |
| Cooper and others | 2017 | Lyme disease and Bell's palsy: an epidemiological study of diagnosis and risk in England |
| Dadvand and others | 2014 | Inequality, green spaces, and pregnant women: roles of ethnicity and individual and neighbourhood socioeconomic status |

| Author | Year | Title |
|---|---|---|
| Foudi and Osés-Eraso | 2022 | Information, experience, and willingness to mitigate mental health consequences from flooding through collective defence |
| Houston and others | 2021 | Social influences on flood preparedness and mitigation measures adopted by people living with flood risk |
| Hunter and others | 2016 | Coastal clustering of HEV; Cornwall, UK |
| Kane and others | 2011 | Understanding occupant heating practices in UK dwellings |
| Kennard and others | 2020 | The associations between thermal variety and health: Implications for space heating energy use |
| Khan and others | 2022 | Association of patient and family reports of hospital safety climate with language proficiency in the US |
| Khieu and others | 2017 | Modelled seasonal influenza mortality shows marked differences in risk by age, sex, ethnicity and socioeconomic position in New Zealand |
| Lewer and others | 2023 | Opioid injection-associated bacterial infections in England, 2002-2021: A time series analysis of seasonal variation and the impact of Coronavirus Disease 2019 |
| Lloyd and others | 2017 | Emergency department presentations of people who are homeless: The role of occupational therapy |
| Maund and others | 2019 | Wetlands for wellbeing: Piloting a nature-based health intervention for the management of anxiety and depression |
| Milojevic and others | 2016 | Methods to estimate acclimatization to urban heat island effects on heat- and cold-related mortality |
| Mutz and others | 2021 | Exploring health in the UK Biobank: associations with sociodemographic characteristics, psychosocial factors, lifestyle and environmental exposures |
| Ogbebor and others | 2018 | Seasonal variation in mortality secondary to acute myocardial infarction in England and Wales: a secondary data analysis |
| Pattnaik | 2023 | Role of health equity in the climate action plans of London boroughs: A health policy report |
| Paudyal and others | 2021 | Clinical characteristics, attendance outcomes and deaths of homeless persons in the emergency department: implications for primary health care and community prevention programmes |
| Pearce and others | 2010 | Environmental justice and health: the implications of the socio-spatial distribution of multiple environmental deprivation for health inequalities in the United Kingdom |
| Rees and others | 2017 | Trend analysis of imported malaria in London; observational study 2000 to 2014 |
| Shi and others | 2024 | Risk of winter hospitalisation and death from acute respiratory infections in Scotland: national retrospective cohort study |
| Wagner and others | 2014 | Migrant health and infectious diseases in the UK: findings from the last 10 years of surveillance |

| Author | Year | Title |
|---|---|---|
| Warner and others | 2023 | Infections in travellers returning to the UK: a retrospective analysis (2015-2020) |

**Exclusion reason: wrong outcome (n=73)**

| Author | Year | Title |
|---|---|---|
| Abourashed and others | 2024 | Development and validation of the MosquitoWise survey to assess perceptions towards mosquitoes and mosquito-borne viruses in Europe |
| Alves and others | 2020 | Climate change policies and agendas: Facing implementation challenges and guiding responses |
| Arnell and others | 2021 | The effect of climate change on indicators of fire danger in the UK |
| Barrett and others | 2022 | Eat or heat: fuel poverty and childhood respiratory health |
| Boeckmann and Zeeb | 2014 | Using a social justice and health framework to assess European climate change adaptation strategies |
| Brooks and others | 2015 | Case studies of community resilience to climate change |
| Butler and others | 2018 | Narratives of recovery after floods: Mental health, institutions, and intervention |
| Caan | 2023 | Poor housing is a problem in heatwaves as well as cold weather |
| Chalabi and others | 2016 | Evaluation of the cold weather plan for England: modelling of cost-effectiveness |
| Climate Just | | Case study: Combining climate change adaptation and social housing refurbishment in the London Borough of Barking and Dagenham |
| Climate Just | | Case study: Health promotion and climate mitigation in Middlesbrough |
| Connon | 2017 | Extreme weather, complex spaces and diverse rural places: An intra-community scale analysis of responses to storm events in rural Scotland, UK |
| Demski and others | 2017 | Experience of extreme weather affects climate change mitigation and adaptation responses |
| Dittrich and others | 2019 | A cost-benefit analysis of afforestation as a climate change adaptation measure to reduce flood risk |
| Fazey and others | 2017 | Community resilience to climate change: Outcomes of the Scottish Borders climate resilient communities project |
| Ferranti and others | 2023 | Incorporating heat vulnerability into local authority decision making: An open access approach |
| Ferlie | 2015 | Commentary on text of interview with Professor Lord Ara Darzi: 'Desirable? Yes; but is it achievable?' |
| Fernandez-Bilbao and others | 2011 | Impacts of climate change on disadvantaged UK coastal communities |

| Author | Year | Title |
|---|---|---|
| Fielding | 2012 | Inequalities in exposure and awareness of flood risk in England and Wales |
| Fielding | 2018 | Flood risk and inequalities between ethnic groups in the floodplains of England and Wales |
| Fragkos and others | 2021 | Equity implications of climate policy: Assessing the social and distributional impacts of emission reduction targets in the European Union |
| Garbutt | 2015 | Assessment of social vulnerability under 3 flood scenarios using an open source vulnerability index |
| Garrett and others | 2023 | Visiting nature is associated with lower socioeconomic inequalities in well-being in Wales |
| Georgiou and others | 2022 | A population-based retrospective study of the modifying effect of urban blue space on the impact of socioeconomic deprivation on mental health, 2009-2018 |
| Gralepois and others | 2016 | Is flood defense changing in nature? Shifts in the flood defense strategy in six European countries |
| Grover and Daniels | 2017 | Social equity issues in the distribution of feed-in tariff policy benefits: A cross sectional analysis from England and Wales using spatial census and policy data |
| Gustin and others | 2018 | Forecasting indoor temperatures during heatwaves using time series models |
| Hart and others | 2018 | Contextual correlates of happiness in European adults |
| Hino and Hall | 2017 | Real options analysis of adaptation to changing flood risk: structural and nonstructural measures |
| Houston and others | 2021 | Environmental vulnerability and resilience: Social differentiation in short- and long-term flood impacts |
| Huaccha | 2023 | Regional persistence of the energy efficiency gap: Evidence from England and Wales |
| Hyland and Donnelly | 2015 | Air pollution and health – the views of policy makers, planners, public and private sector on barriers and incentives for change |
| Jones and Mays | 2016 | The experience of potentially vulnerable people during cold weather: implications for policy and practice |
| Kantamaneni | 2019 | Evaluation of social vulnerability to natural hazards: a case of Barton on Sea, England |
| Kaźmierczak and Cavan | 2011 | Surface water flooding risk to urban communities: Analysis of vulnerability, hazard and exposure |
| Kennedy-Asser and others | 2022 | Projected risks associated with heat stress in the UK Climate Projections (UKCP18) |
| Khare and others | 2015 | Heat protection behaviour in the UK: results of an online survey after the 2013 heatwave |

| Author | Year | Title |
|---|---|---|
| Kidd and others | 2021 | The climate change-homelessness nexus |
| Kidd and others | 2023 | A response framework for addressing the risks of climate change for homeless populations |
| Kim and Donohue | 2011 | Demographic, developmental and life-history variation across altitude in Erysimum capitatum |
| Kings and Ilbery | 2010 | The environmental belief systems of organic and conventional farmers: Evidence from central-southern England |
| Kreslake and others | 2016 | Developing effective communication materials on the health effects of climate change for vulnerable groups: a mixed methods study |
| Lacey-Barnacle | 2020 | Proximities of energy justice: contesting community energy and austerity in England |
| Laszkiewicz and others | 2023 | Who does not use urban green spaces and why? Insights from a comparative study of thirty-three European countries |
| leBrasseur | 2023 | Citizen sensing within urban greenspaces: Exploring human wellbeing interactions in deprived communities of Glasgow |
| Lomas and others | 2021 | Dwelling and household characteristics' influence on reported and measured summertime overheating: A glimpse of a mild climate in the 2050's |
| Macintyre and others | 2018 | Assessing urban population vulnerability and environmental risks across an urban area during heatwaves – implications for health protection |
| Majekodunmi and others | 2020 | A spatial exploration of deprivation and green infrastructure ecosystem services within Glasgow city |
| Munro and others | 2017 | Effect of evacuation and displacement on the association between flooding and mental health outcomes: a cross-sectional analysis of UK survey data |
| Olsen and others | 2023 | Inequalities in neighbourhood features within children's 20-minute neighbourhoods and variation in time spent locally, measured using GPS |
| Paauw and others | 2024 | Recognition of differences in the capacity to deal with floods-A cross-country comparison of flood risk management |
| Parker | 2023 | Barriers to green inhaler prescribing: ethical issues in environmentally sustainable clinical practice |
| Petzold | 2016 | Limitations and opportunities of social capital for adaptation to climate change: a case study on the Isles of Scilly |
| Preston and others | 2013 | Distribution of carbon emissions in the UK : implications for domestic energy policy |
| Reynolds and others | 2019 | Healthy and sustainable diets that meet greenhouse gas emission reduction targets and are affordable for different income groups in the UK |

| Author | Year | Title |
|---|---|---|
| Rundblad and others | 2010 | Communication, perception and behaviour during a natural disaster involving a 'Do Not Drink' and a subsequent 'Boil Water' notice: a postal questionnaire study |
| Sanchez-Guevara and others | 2019 | Assessing population vulnerability towards summer energy poverty: Case studies of Madrid and London |
| Sanderson and Ford | 2017 | Projections of severe heat waves in the United Kingdom |
| Sayers and others | 2015 | Climate Change Risk Assessment 2017: Projections of future flood risk in the UK |
| Scheelbeek and others | 2020 | United Kingdom's fruit and vegetable supply is increasingly dependent on imports from climate-vulnerable producing countries |
| Schulte and Hudson | 2023 | A cross-sectional study of inequalities in digital air pollution information access and exposure reducing behavior uptake in the UK |
| Sheng and others | 2023 | Climate shocks and wealth inequality in the UK: evidence from monthly data |
| Shikder and others | 2012 | Summertime impact of climate change on multi-occupancy British dwellings |
| Shoari and others | 2022 | Towards healthy school neighbourhoods: A baseline analysis in Greater London |
| Soetanto and others | 2017 | The perceptions of social responsibility for community resilience to flooding: the impact of past experience, age, gender and ethnicity |
| Tang and Rundblad | 2015 | The potential impact of directionality, colour perceptions and cultural associations on disaster messages during heatwaves in the UK |
| Town and Country Planning Association | 2016 | Planning for the climate challenge? Understanding the performance of English local plans |
| Vellei and others | 2017 | Overheating in vulnerable and non-vulnerable households |
| Wade and others | 2015 | Developing H++ climate change scenarios for heat waves, droughts, floods, windstorms and cold snaps |
| Watkiss and others | 2016 | Climate change impacts on the future cost of living (SSC/CCC004) |
| Wolf and others | 2010 | Heat waves and cold spells: an analysis of policy response and perceptions of vulnerable populations in the UK |
| Zahiri and Gupta | 2023 | Examining the risk of summertime overheating in UK social housing dwellings retrofitted with heat pumps |
| Zhou and others | 2023 | Exploring socio-ecological inequalities in heat by multiple and composite greenness metrics: A case study in Belfast, UK |

**Exclusion reason: duplicate reference (n=16)**

| Author | Year | Title |
|---|---|---|
| Arbuthnott and others | 2020 | Years of life lost and mortality due to heat and cold in the three largest English cities |
| Bryan and others | 2020 | The health and well-being effects of drought: assessing multi-stakeholder perspectives through narratives from the UK |
| Dear and McMichael | 2011 | The health impacts of cold homes and fuel poverty |
| Health Protection Agency | 2012 | Health effects of climate change in the UK 2012: current evidence, recommendations and research gaps |
| Hocking and others | 2020 | Heat, energy efficiency, smart technology and health: A review of evidence from high-income countries, with a focus on the UK |
| Karamanos and others | 2021 | Air pollution and trajectories of adolescent conduct problems: the roles of ethnicity and racism; evidence from the DASH longitudinal study |
| Konstantinoudis and others | 2021 | Ambient heat exposure and COPD hospitalisations in England: A nationwide case-crossover study during 2007-2018 |
| Milojevic and others | 2016 | Methods to estimate acclimatization to urban heat island effects on heat- and cold-related mortality |
| Murage and others | 2018 | Variation in cold-related mortality in England since the introduction of the Cold Weather Plan: Which areas have the greatest unmet needs? |
| Oyebanjo and Bushell | 2014 | A critical evaluation of the UK SunSmart campaign and its relevance to Black and minority ethnic communities |
| Seklecka and others | 2017 | Mortality effects of temperature changes in the United Kingdom |
| Tainio and others | 2017 | Mortality, greenhouse gas emissions and consumer cost impacts of combined diet and physical activity scenarios: a health impact assessment study |
| Taylor and others | 2021 | Projecting the impacts of housing on temperature-related mortality in London during typical future years |
| Tieges and others | 2020 | The impact of regeneration and climate adaptations of urban green-blue assets on all-cause mortality: A 17-year longitudinal study |
| Wolf and others | 2010 | Heat waves and cold spells: an analysis of policy response and perceptions of vulnerable populations in the UK |
| Zafeiratou and others | 2023 | Assessing heat effects on respiratory mortality and location characteristics as modifiers of heat effects at a small area scale in Central-Northern Europe |
